# Joint modeling HIV and HPV using a new hybrid agent-based network and compartmental simulation technique

**DOI:** 10.1101/2022.12.25.22283941

**Authors:** Xinmeng Zhao, Chaitra Gopalappa

## Abstract

**Introduction:** Persons living with HIV have a disproportionately higher burden of HPV-related cancers. Causal factors include both behavioral and biological. While pharmaceutical and care support interventions help address biological risk of coinfection, as social conditions are common drivers of behaviors, structural interventions are key part of behavioral interventions. Joint modeling sexually transmitted diseases (STD) can help evaluate optimal intervention combinations for overall disease prevention. While compartmental modeling is sufficient for faster spreading HPV, network modeling is suitable for slower spreading HIV. However, using network modeling for jointly modeling HIV and HPV can generate computational complexities given their vastly varying disease epidemiology and disease burden across sub-population groups.

**Methods:** We applied a recently developed mixed agent-based compartmental (MAC) simulation technique, which simulates persons with at least one slower spreading disease and their immediate contacts as agents in a network, and all other persons including those with faster spreading diseases in a compartmental model, with an evolving contact network algorithm maintaining the dynamics between the two models. We simulated HIV and HPV in the U.S. among heterosexual female, heterosexual male, and men who have sex with men (men only and men and women) (MSM), sub-populations that mix but have varying HIV burden, and cervical cancer among women. We conducted numerical analyses to evaluate the contribution of behavioral and biological factors to risk of cervical cancer among women with HIV.

**Results:** The model outputs for HIV, HPV, and cervical cancer compared well with surveillance estimates. Behavioral factors significantly contributed to risk of HIV-HPV co-infection, and biological factors further exacerbated cancer burden among persons with HIV, with the fraction attributed to each factor sensitive to disease burden.

**Conclusions:** This work serves as proof-of-concept of the MAC simulation technique for joint modeling related diseases with varying epidemiology in sub-populations with varying disease burden. Future work can expand the model to simulate sexual and care behaviors as functions of social conditions, and further, jointly evaluate behavioral, structural, and pharmaceutical interventions for overall STD prevention.

## Introduction

Human papillomavirus virus (HPV) is one of the most common sexually transmitted diseases (STDs) in the United States, about 80% of sexually active persons are estimated to have acquired HPV at least once during a lifetime [1,2]. Although most HPV infections resolve on their own within 2 years, persistent infection with certain types of HPV (high-risk or oncogenic HPV) are a causal factor for most cases of cervical cancer [3–5] and anal cancers [6–8], and a likely cause of substantial proportions of other genital neoplasms and oral squamous cell carcinomas [4,9]. On average, each year in the United States, about 12,000 women are estimated to be diagnosed with cervical cancer, and an additional 11,000 women and 16,000 men are annually diagnosed with other HPV-associated cancers [10]. Persons with impaired cell-mediated immunity, such as persons with human immunodeficiency virus (HIV) infection, particularly suffer from increased risk of HPV infection and subsequent genital tract neoplasias and cancers [11–13]. For example, the risk of HPV infection and cervical cancer incidence among women with HIV compared to women without HIV were estimated to be about ∼2-6 times [14–16] and ∼2-7 times higher, respectively [17–19].

Causal factors for higher risk of HPV infection among persons with HIV could be attributed to behavioral factors, i.e., individual sexual behaviors, partnership networks, and care behaviors, and biological factors, i.e., a compromised immune system from HIV can biologically increase the risk of other diseases [20,21]. Studies in the literature have investigated the attribution to biological factors by using observational data and statistically accounting for the confounding factors of sexual behavior. These include estimating associations between HIV exposure and HPV acquisition [22–25], rate of HPV clearance [26–28], and risk of HIV on disease progression and cervical cancer pathology [29–35]. The primary focus of these studies is on biological factors, and most studies are based on statistical methods such as multivariate regression analyses. However, they do not consider the dynamics of behavioral factors, such as changes in individual behaviors, partnership networks, and system-level changes in care.

Quantitative estimations of the risk of HIV-HPV coinfection, attributable to each type of factor, behavioral and biological, could help inform the type of intervention needs. Specifically, social conditions, such as poverty, unemployment, stigma, and discrimination, are key drivers of behaviors that increase risk of STIs, e.g., higher number of partners, higher condomless sex, lower care uptake among persons experiencing homelessness than among those with stable housing [36–39]. Consequently, though the prevalence of HIV in the U.S. is low, it is concentrated among the most vulnerable populations. Among persons living with diagnosed HIV infection, an estimated 44% had a disability (including physical, mental, and emotional disabilities), 41% were unemployed, 43% had household incomes at or below the federal poverty threshold, and 10% were experiencing homelessness [40,41]. Thus, structural interventions, such as health care coverage, subsidized housing and food programs, and access to mental healthcare are key part of behavioral interventions for prevention of STIs [42–45]. On the other hand, biological risk of coinfection would additionally require pharmaceutical and care support programs for disease management.

A dynamic model of sexual transmission networks that jointly simulates HIV and HPV will serve as a suitable decision-analytic tool for analyses of structural and disease-specific interventions. Several models in the literature have jointly simulated HIV and HPV for economic analyses of interventions such as antiretroviral therapy treatment and pre-exposure prophylaxis for HIV and HPV vaccinations and cervical cancer screening for women [46–50]. Most models are deterministic compartmental models [46,47,50], and the models that are individual based do not include a transmission model [48] or do not explicitly simulate partnership networks [49], which limit the capacity to fully account for the behavioral interactions between HIV and HPV infections.

We built a joint HIV-HPV simulation model using a mixed agent-based network and compartmental (MAC) simulation framework and conducted numerical analyses to evaluate the risk of coinfection attributable to behavioral and biological factors. These numerical findings will help inform intervention needs and, in future work, help conduct model evaluations of both structural interventions and disease-specific interventions. While observational studies can help estimate the costs, the behavioral interactions in the model can help evaluate the impact of structural interventions on overall STI prevention.

## Methods

MAC is a recently developed simulation framework for joint modeling diseases of varying epidemiology [51]. While compartmental modeling is sufficient for higher prevalence diseases such as HPV, Chlamydia, and Gonorrhea, agent-based network modeling is preferred for slower spreading diseases such as HIV and Hepatitis C, as the network structures have a larger influence on disease spread [52]. However, agent-based modeling alone will be computationally challenging for national joint disease modeling. For example, in the U.S., HIV prevalence is about 0.4% and 25% of HIV infected persons are women [53] and cervical cancer prevalence among HIV infected women is 0.3% [54]. Thus, simulating 100,000 persons representative of the U.S. population will generate 100 women with HIV, and no cases of cervical cancer. Increasing the number of samples will exponentially increase the challenges with computational tractability. Similar challenges are faced when modeling sub-populations with disproportionately varying disease burden. For example, in the U.S., about 64% of HIV cases are among men who have sex with men [53], who constitute about 2% of the U.S. population [55]. Most HIV network models in the literature simulate sub-populations separately, which overlooks the mixing associated between sub-groups, e.g., about half of new HIV cases among women were linked to transmissions from MSM [56,57]. The MAC framework overcomes the computational challenges by simulating persons with at least one lower prevalence disease and their immediate contacts in an agent-based network model, and all other persons including those with only higher prevalence diseases in a compartmental model, using an agent-based evolving network algorithm (ABENM) to maintain the network dynamics between persons in the two models [58,59]. The MAC simulation framework has been described elsewhere [51], and ABENM for simulating sexual transmission networks in the U.S. has been applied to the ‘Progression and Transmission of HIV’ (PATH 4.0) model in the U.S. [59], and extensively validated against multiple epidemic and network metrics from the U.S. National HIV Surveillance Systems.

In this work, we used MAC to build a two-disease model, calibrating to lower prevalence HIV and higher prevalence HPV, and validating against data in the U.S., to serve as proof-of-concept of the MAC framework. We provide a brief overview of MAC below and the HIV-HPV model development and calibration.

### Overview of MAC

We present an overview of the MAC simulation framework for HIV-HPV modeling in Fig 1. All persons in the population are ether in the network or in the compartmental model. Persons infected with the lower prevalence HIV (they may also be infected with HPV), and their immediate contacts are tracked in the network and all other persons, including those with only higher prevalence HPV and those uninfected with either disease are tracked in a compartmental model. We newly calibrated an HPV model for the U.S. population and adopted the HIV model from the previously validated PATH 4.0 model [59]. Details of MAC are presented in [51], we present below an overview of the computational structure of MAC, an overview of the calibration of HIV and HPV, and numerical analyses. We simulated HIV and HPV among three transmission risk groups: heterosexual women (HETF), heterosexual men (HETM), and men who have sex with men (MSM) (men only and women). We present in the Appendix more details of the computational structure of MAC in Appendix S1, including HIV and HPV modules in Appendix S1.3, and details of the HPV model calibration in Appendix S2.

**Fig 1.**
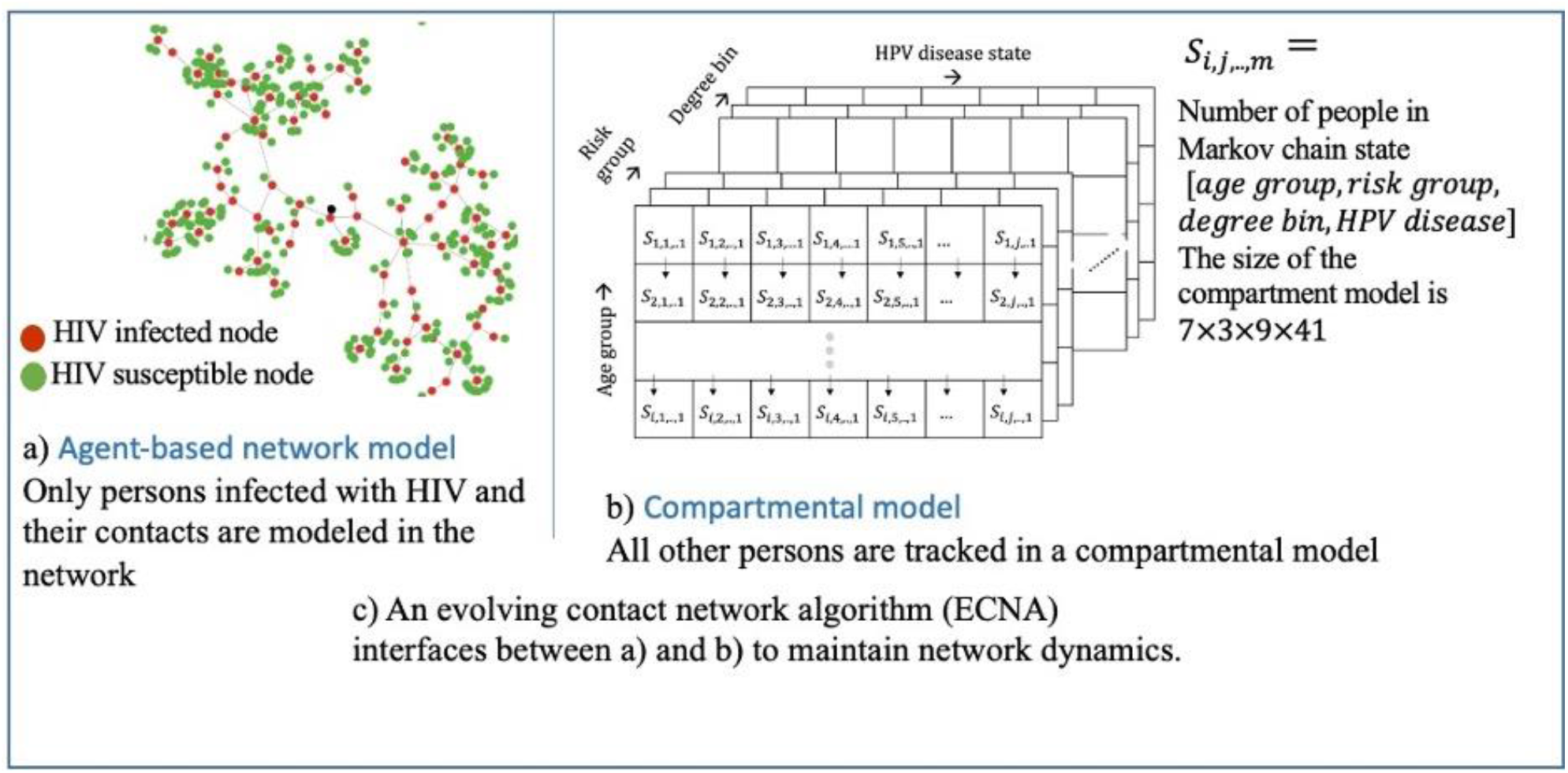
Computational structure of MAC simulation framework for multi-disease modeling: a) HIV, HPV, and b) associated cancer.

#### Computational structure of MAC

The model tracks HIV-infected persons and immediate contacts using a dynamic graph *G*_*t*_(𝒩, ℰ), with the number of nodes in the graph *Q*_*t* =_ |𝒩(*G*_*t*_)| and the number of edges |ℰ(*G*_*t*_)| dynamically changing over time *t* as persons become newly infected with HIV and their immediate contacts are added to the network. Each person in the network has attributes such as age, transmission-group, degree (number of lifetime partnerships), geographic jurisdiction, HIV-related disease and care continuum stages, and HPV related disease and care stages.

The model tracks all other persons in a compartment model, using an array *S*_*t*_ of size *A* × *R* × *D* × 𝒢 × *H*, where, *A* is the number of age-groups, *R* is the number of transmission-groups, *D* is the number of degree-bins (degree is the number of lifetime partners per person, degrees are grouped into bins analogous to age grouped into age-groups), 𝒢 is the number of geographic jurisdictions (in our numerical analyses we assumed 𝒢 = 1, corresponding to a national jurisdiction), and *H* is the number of health states related to HPV including disease-free state. Each element of the array 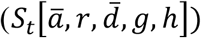 is the number of people in that specific category. We use a dash for age-group and degree-bin notations to indicate that they are grouped intervals in the compartmental model, unlike in the network where each node has a discrete value. A summary list of notations is presented in Appendix Table S1.

Thus, 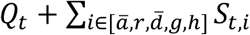, would be the total number of people in the population at time *t*. As all HIV infected persons and exposed partners are in the network, HIV transmissions and disease stage progressions are modeled at the individual level (as typically done in agent-based modeling). HPV transmissions and progression are modeled using differential equations (as typically done in compartmental modeling technique) but with the consideration that people in both the network *G*_*t*_(𝒩, ℰ) and compartmental array *S*_*t*_ can be infected with HPV.

The model adopts the concepts from a previously developed ABENM simulation technique [58,59] to maintain the dynamics between the network *G*_*t*_(𝒩, ℰ) and compartmental array *S*_*t*_, including transitioning people from *S*_*t*_ to *G*_*t*_(𝒩, ℰ) when they become newly infected with HIV. As HIV infection is chronic once persons enter the network *G*_*t*_(𝒩, ℰ) they do not transition back to *S*_*t*_. More details of the MAC computational structure are provided in Appendix S1.1 and S1.2. The overall epidemiological, demographical, and network dynamics are maintained through simulation of four main modules that are run at every time-step (monthly) of the simulation: compartmental module for HPV transmissions and progression, network transmission module for HIV transmissions, ECNA network generation module for maintaining partnership dynamics and network dynamics between the compartmental model and the network, and network disease progression module for HIV progression. We discuss each module in Appendix S1.3 and provide an overview of each below.

The compartmental module updates the demographic features (births, aging, and deaths) among persons tracked through the array *S*_*t*_. It also models transmission and progression features of high-prevalence diseases (HPV here) among persons tracked through the array *S*_*t*_ and in the network *G*_*t*_(𝒩, ℰ). Specifically, as in typical compartment modeling, it uses difference equations to calculate the rate of transitioning between compartments that represent HPV disease stages. It also uses those rates to determine the number of people in the network to transition between states corresponding to those compartments. A key feature here is approximating network structures within the compartmental model by using degree-bins as part of the model framework (a dimension in array *S*_*t*_) and using a degree-mixing matrix to simulate partnerships that can occur with people in different degree-bins. Degree, in addition to age and transmission-group, between partners are correlated [60] and thus this feature helps better capture the network dynamics even in the compartmental model. While the distributions for partnership mixing by age-group, transmission-group, and degree-bin are applied at the individual level in the network they are applied at the aggregated level in the compartmental model. The general MAC computational structure of the compartmental module is presented in Appendix S1.3.1 and its application specific to HPV is presented in Appendix S2.

The network transmission module uses an individual-level Bernoulli transmission equation to determine if nodes in the network *G*_*t*_(𝒩, ℰ) exposed to a lower prevalence disease (HIV here) become infected. Transmissions are determined at the individual level using the network structure and individual-level sexual behaviors and transmission risk factors. We present an overview of the HIV Bernoulli transmission equation in Appendix 1.3.2. Note that, as persons in the compartmental model are not partners of any person infected with the lower prevalence diseases (HIV here), their chance of infection is zero. Further note that persons can move from the compartmental model to the network (Fig 1) upon becoming partners of an HIV-infected person, modeled using the ECNA module (discussed below), which would then expose them to the infection. We adopted the HIV transmission module from PATH 4.0 [59].

The ECNA module controls the overall network dynamics of partnerships between persons in the compartmental model and the network. Specifically, for every node newly infected with HIV in the network, it determines the number of new partnerships to generate and the features of each of those new partners, including their number of lifetime partners, their transmission-group, and their current age-group. The module then randomly selects susceptible persons who meet these criteria and moves them from the compartmental model to the network. As all life-time partnerships of an HIV-infected person are in the network, contacts need to be activated and deactivated as per when the partnership initiates and terminates. The ECNA module determines these partnership details, such as the age of both partners and simulation times at partnership initiation and termination. It does so by using multiple sub-algorithms developed using concepts from machine learning, stochastic processes, and optimization, which are presented in [58,59] and are summarized in Appendix S1.3.3.

Finally, the network disease progression module updates the individual-level demographic and disease dynamics for every person infected with HIV (and other lower prevalence diseases in its general application) in the network. We adopted the HIV progression module from [56,59].

### Model calibration and validation

#### HIV

We adopted the validated HIV model from PATH 4.0 as lower prevalence Disease 1 [59]. PATH 4.0 was validated to match well against data from the National HIV Surveillance System (NHSS) for both epidemic features and network features. Details of the ABENM, PATH 4.0, and its validation are presented in [59]; we give a brief description below. PATH 4.0 simulates sexual transmission of HIV in the U.S. in three transmission risk groups: HETF, HETM and MSM. The model is first initialized to be representative of people living with HIV (PWH) in the U.S. in 2006, using data from several studies. These include demographical, sexual behavioral, clinical, and HIV care and treatment behavioral studies that originated from multiple large national surveillance and survey systems in the U.S., along with other small studies. The surveillance and survey systems include the NHSS, the Medical Monitoring Project (MMP), the HIV Outpatient Study (HOPS), the National HIV Behavioral Surveillance (NHBS), the National Survey for Family Growth (NSFG), and the National Survey for Sexual Health and Behavior (NSSHB) [61–66]. After initialization to 2006, the model then simulates HIV from 2006 to 2017 in monthly-time steps, using data for annual changes in care continuum from NHSS [59].

#### HPV

We constructed a new HPV and cervical cancer model as higher prevalence Disease 2. Detailed description of model development, data assumptions, calibration, and validation are presented in Appendix 2, and summarized below. The compartmental model array *S*_*t*_ was of size 7 × 3 × 9 × 1 × 41, corresponding to size of age-group, transmission risk group, degree-bin, geographical jurisdiction, and HPV health state. Here we simulated an overall national population, thus, there was only 1 jurisdiction.

Note that, as in the HIV model, there were three transmission-groups, HETF, HETM, and MSM. For HETF, we simulated HPV and cervical cancer. For HETM, we simulated only HPV as the risk of sequelae among HETM is low. For MSM, anal HPV can lead to cancers such as anal cancer, which has a similar epidemiology as cervical cancer [67]. We did not specifically model anal cancer. However, as HIV prevalence and sexual networks among MSM are significantly different than among heterosexuals, to understand the mathematical sensitivity of these network dynamics on metrics of interest (see numerical analyses below), we used the disease progression data for HPV and cervical cancer to model HPV and anal cancer among MSM. Thus, we validated this model for only HPV and cervical cancer among women.

Sexual behavioral data, such as the number of sex acts, condom use, and partnership mixing across age-groups and transmission groups, were kept consistent between the compartmental and network models (as they collectively represent the U.S population). That is, data specific to age-group, transmission risk group, and degree-bin from PATH 4.0 were also used in the estimation of HPV infection rates, i.e., the rate of transitioning from susceptible to first stage of infection. Data for the state-transitions related to natural HPV progression and regression were gathered from literature studies [47,68–70]. We calibrated the per-act probability of transmission specific to HPV-genotype by fitting the natural history model to common calibration targets e.g., high-risk HPV genotype frequency among cervical intraepithelial neoplasia (CIN) 1, 2, 3, and normal cytology and age-and genotype-specific high-risk HPV prevalence among normal cytology [71] as typically done in other models in the literature [72]. Results show a good fit to most metrics (Fig 2). To assess the validity of the model, we compared pre-screening cervical cancer incidence and mortality from our model with the data obtained from the Connecticut Tumor Registry (Fig 3a) [73].

**Fig 2.**
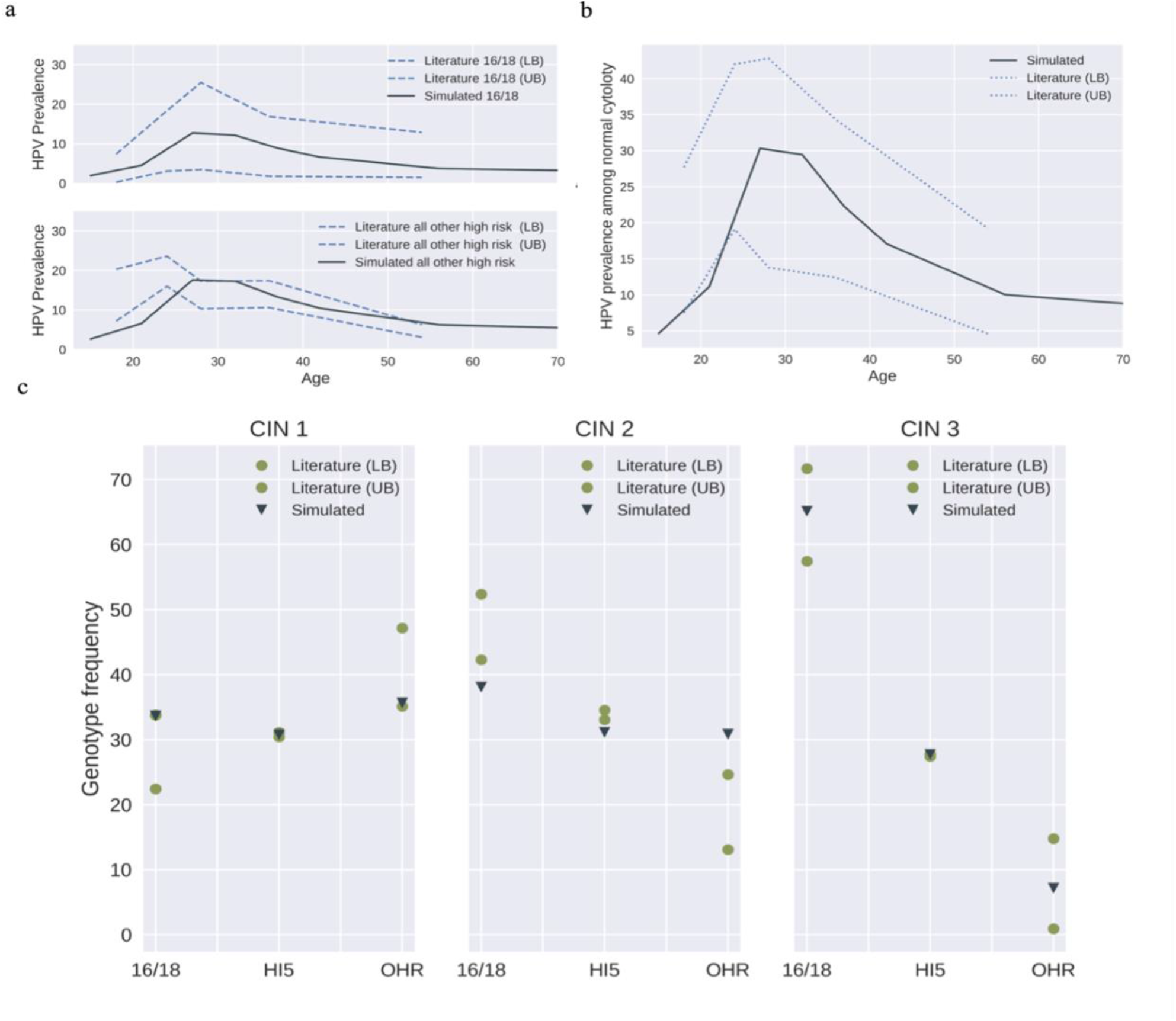
HPV and cervical cancer natural history model calibration results. a) Age-and genotype-specific high-risk HPV prevalence among normal cytology; b) Age-specific HPV prevalence among normal cytology; c) High-risk HPV genotype frequency among cervical intraepithelial neoplasia (CIN) 1, 2 and 3. (Data source: [71]). 16/18 represent HPV-16/18 pooled, HI5 represents HPV-31/33/45/52/58 pooled, while OHR represents other high-risk genotypes except HPV-16/18/31/33/45/52/58.

**Fig 3.**
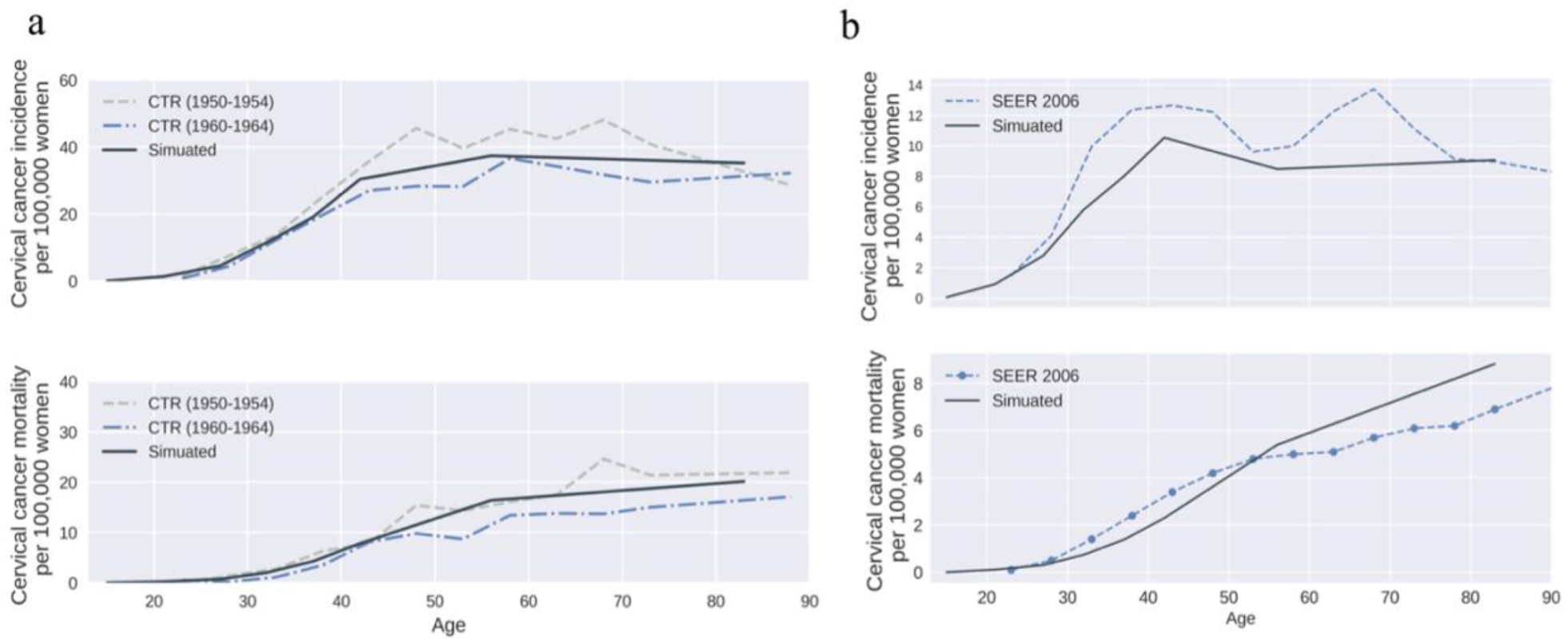
HPV and cervical cancer natural history model validation results. a) Age-specific cervical cancer incidence and mortality per 100,000 women (without screening); b) Age-specific cervical cancer incidence and mortality per 100,000 women (with screening). (Data source: a) [73]; b) SEER 2006 [76]).

We used the calibrated model to simulate and validate HPV in the U.S under cervical cancer screening, using data corresponding to 2006. These data include screening rates, cytology screening sensitivity, proportions receiving follow-up colposcopy/biopsy and proportion receiving precancer treatment for those in pre-cancer stages, and additionally cancer-screening sensitivity for those in cancer stages [74,75]. Cervical cancer incidence and mortality in 2006 with screening [76] compare well with surveillance estimates (Fig 3b). More details on calibration and validation are provided in Appendix 2.5.

### Numerical analyses

We focused the numerical analyses on estimating the differences in HPV disease burden among people with HIV compared to people without HIV. In this regard, we define the following relative risk metrics, relative prevalence of HPV (*RP*_*HPV*_) and relative incidence of cancer (*RI*_*cancer*_).

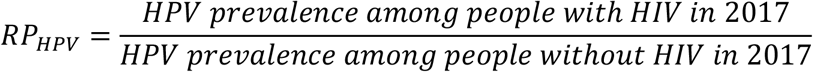

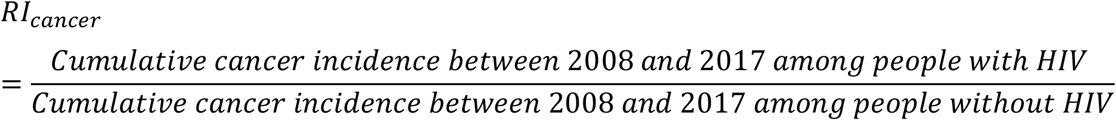

Prevalence is the proportion of people living with the disease, calculated at the end of 2017, and includes both diagnosed and undiagnosed cases. Cancer incidence is the number of newly diagnosed cases in a year. We used prevalence for *RP*_*HPV*_ but cumulative incidence for *RI*_*cancer*_ because persons in an HPV infection state can transition back to a susceptible state, while persons transitioning to a cancer state remain in that state until mortality.

### Scenarios modeled

To determine the attribution of biological and behavioral factors to the higher HPV burden among women with HIV, we estimated *RP*_*HPV*_ and *RI*_*cancer*_ under four assumptions of biological risk (discussed below). Further, to evaluate the sensitivity of *RP*_*HPV*_ and *RI*_*cancer*_ to changes in HPV burden over time, caused by changes in interventions, we evaluated the metrics under three HPV intervention assumptions (discussed below). Thus, we evaluated a combination of twelve scenarios. We discuss each of the four biological risk assumptions and three HPV intervention assumptions below. As HIV care has significantly increased over the past decade, to test the sensitivity of the relative risk metric to HIV burden, for each scenario, we evaluated *RP*_*HPV*_ at two-time points years 2010 and 2017.

#### Biological risk assumptions

- *No biological risk*: Assumes incidence and progression rates for HPV and related cancers would be the same for persons with HIV and without HIV (note: incidence and progression rates do change by other factors such as age-group, sexual behavior, etc.). The values for these were taken from data in the literature and are discussed in Appendix S2.4.1. We assume these to be the basecase rates.
- *Increased risk of disease progressio*n: Assumes higher rates of HPV progression and lower rates of HPV regression for persons with HIV infection compared to persons without (through use of a factor multiplied to basecase rates). Further, the value of the multiplier varied by HIV-disease stage. Data for these multipliers were based on studies in the literature (Appendix Table S2) [47,77,78]. Though these data were specific to cervical cancer, due to lack of data we assumed same multipliers when modeling anal cancer among MSM, as studies suggest higher biological risk for anal cancer [11–13].
- *Increased risk of HPV acquisitio*n: Assumes a higher rate of HPV acquisition for persons with HIV compared to persons without HIV by using a multiplier to the basecase infection rates. Data for these multipliers were based on studies in the literature (Appendix Table S2) [47]. Though these data were specific to cervical cancer, due to lack of data we assumed same multipliers when modeling anal cancer among MSM, as studies also suggest higher biological risk for anal cancer [11–13].
- *All increased ris*k: Combines the above two scenarios by assuming higher rates of HPV acquisition, higher rates of HPV progression, and lower rates of HPV regression for persons with HIV infection compared to persons without.

#### Intervention assumptions

- *Pre-screening*: Assumes symptom-based diagnosis for HPV and cervical cancer.
- *Screening-only*: Assumes screening rates as per year 2006 in the U.S. and keeps it constant over the period 2006 to 2017.
- *Status-quo* intervention: Assumes screening rates as per year 2006 in the U.S. and keeps it constant over the period 2006 to 2017. Additionally, it models HPV vaccinations. It assumes vaccinations initiated in 2007 for females and 2010 for males aged 13-17 years, assumes a quadrivalent vaccine type for the period before 2015 and nonavalent vaccine for 2015 and after, and models’ vaccination rates to vary per year as per data in the U.S. [79–81]. This scenario is the closest representation for screening practice and HPV vaccination coverage in the U.S. for recent years.

### Metrics gathered

We simulated each of the 12 scenarios 10 times (simulating over the period from 2006 to 2017). We present the mean and range of *RP*_*HPV*_ and *RI*_*cancer*_ across the 10 runs. Values of greater than 1 indicate higher burden among persons with HIV compared to persons without. We also estimated the fraction of increased risk attributed to biological factors as [(*RP*_*HPV*_ in ‘all increased risk’ minus *RP*_*HPV*_ in ‘no biological risk’) divided by (*RP*_*HPV*_ in ‘no biological risk’ minus 1)]. To better interpret observed values of *RP*_*HPV*_ and *RI*_*cancer*_, we also extracted results for the following metrics related to network and epidemic dynamics. Average degree (number of lifetime partners) among HIV+ (*d*_*HIV*+_), average degree among HPV+ (*d*_*HPV*+_), average degree among overall population (Overall) (*d*_*overall*_), HPV prevalence among HIV+, HPV prevalence among HIV-, cancer incidence among HIV+, and cancer incidence among HIV-. We extracted these metrics specific to each transmission risk group, HETF, HETM, and MSM.

## Results

As expected, in all scenarios, HIV prevalence was highest among MSM (∼8%), moderate among HETF (∼0.12%), and lowest among HETM (∼0.06%), which matches with estimates from the U.S. National HIV Surveillance System [82]. Considering the lower prevalence of HIV among HETF, the overall HPV prevalence and cervical cancer incidence among women did not significantly vary across the biological risk assumptions (though values varied among HIV+ women as discussed later). Overall rates of HPV prevalence were 19%, 25%, and 29% among HETF, HETM, and MSM, respectively. Annual rates of cervical cancer incidence (and mortality), among HETF, in pre-screening, screening-only, and status-quo scenarios were 23 (10), 7 (3), and 7 (3) per 100,000 persons, respectively. There were no differences between screening-only and status-quo given the low vaccination uptakes and short timeline from vaccine introduction. These results match with surveillance data, which report an incidence of 7.6 per 100,000 persons and mortality rate of 2.4 per 100,000 persons in 2006, the year prior to introduction of vaccines, and an incidence of 6.7 per 100,000 persons and mortality rate of 2.2 per 100,000 persons by the end of 2017 [76].

In the ‘status-quo, no biological risk’ scenario, *RP*_*HPV*_ was significantly greater than 1 in all three transmission-groups, however, it was highest among HETF (1.41(1.36-1.44)) (Table 1c) followed by MSM (1.36(1.34-1.38)) (Table 3c), and closer to 1 for HETM (1.13(1.01-1.16)) (Table 2c). To recollect, overall HPV prevalence was highest among MSM and HETM, and overall HIV prevalence was high among MSM, moderate among HETF, and low among HETM. Thus, there was no consistent pattern when comparing differences in *RP*_*HPV*_ across transmission risk-group with HIV prevalence or HPV prevalence alone.

**Table 1a:** Numerical analyses of HIV and HPV disease burden and relevant network metrics under “prescreening” assumption (HETF)

|  | Prescreening |  |  |  |
| --- | --- | --- | --- | --- |
|  | No biological risk | All increased risk | Increased risk of progression | Increased risk of HPV acquisition |
| Average degree for HIV+ | 23.08(21.24-24.13) | 23.29(22-24.42) | 23.25(22.05-24.41) | 23.25(21.6-24.61) |
| Average degree for overall population | 13.39(13.38-13.39) | 13.39(13.39-13.39) | 13.39(13.39-13.39) | 13.39(13.38-13.39) |
| Average degree for HPV+ | 19.13(19.12-19.13) | 19.13(19.12-19.14) | 19.13(19.12-19.14) | 19.13(19.12-19.14) |
| Average degree or HIV+ and HPV+ | 29.03(27.19-32.52) | 29.57(26.27-31.99) | 30.03(27.31-32.3) | 29.13(26.28-31.13) |
| HPV prevalence among total population (2010) | 0.21(0.21-0.21) | 0.21(0.21-0.21) | 0.21(0.21-0.21) | 0.21(0.21-0.21) |
| HPV prevalence among total population (2017) | 0.2(0.2-0.2) | 0.2(0.2-0.2) | 0.2(0.2-0.2) | 0.2(0.2-0.2) |
| Relative risk (HPV prevalence, 2010) | 1.29(1.26-1.33) | 1.4(1.32-1.48) | 1.32(1.27-1.36) | 1.36(1.29-1.39) |
| Relative risk (HPV prevalence, 2017) | 1.35(1.27-1.45) *^ | 1.58(1.51-1.64) *+^ | 1.44(1.35-1.55) *+^ | 1.48(1.43-1.51) *+^ |
| Cumulative new cancer incidence among HIV+ over year 2008-2017 (per 100, 000 persons) | 359.07(138.64-567.37) | 1374.64(979.54-1806.82) | 1420.04(877.43-1852.22) | 475.71(257.81-766.67) |
| Cumulative new cancer incidence among HIV- over year 2008-2017 (per 100, 000 persons) | 287.63(287.59-287.67) | 287.65(287.59-287.69) | 287.64(287.59-287.68) | 287.64(287.59-287.67) |
| Relative risk (cumulative cancer incidence over year 2008-2017) | 1.25(0.48-1.97) | 4.78(3.41-6.28) *+ | 4.94(3.05-6.44) *+ | 1.65(0.9-2.67) |

\* indicates significant compared to relative risk = 1
+ indicates significant compared to “no biological risk” within same intervention assumption
^ indicates significant compared to 2010 estimates where HIV burden was lower
† indicates significant compared to “prescreening”
The criterion for significance is $p < 0.05$ .

**Table 1b:**
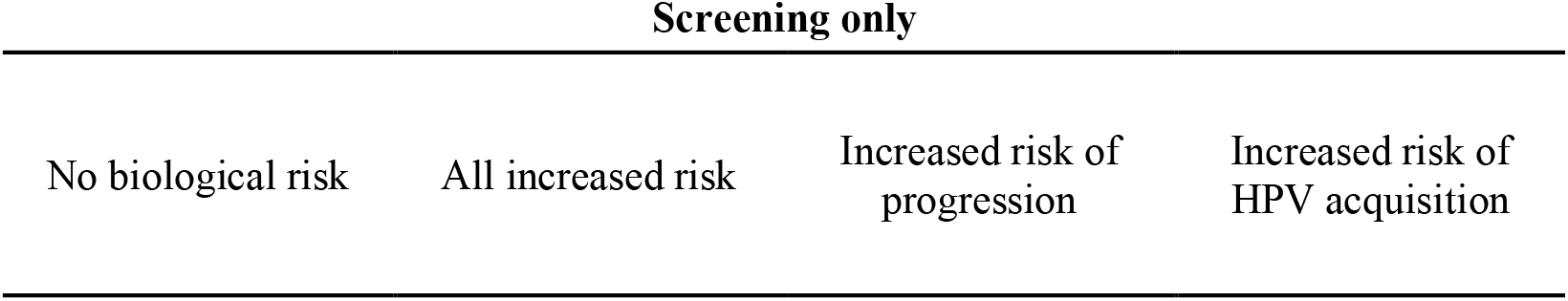
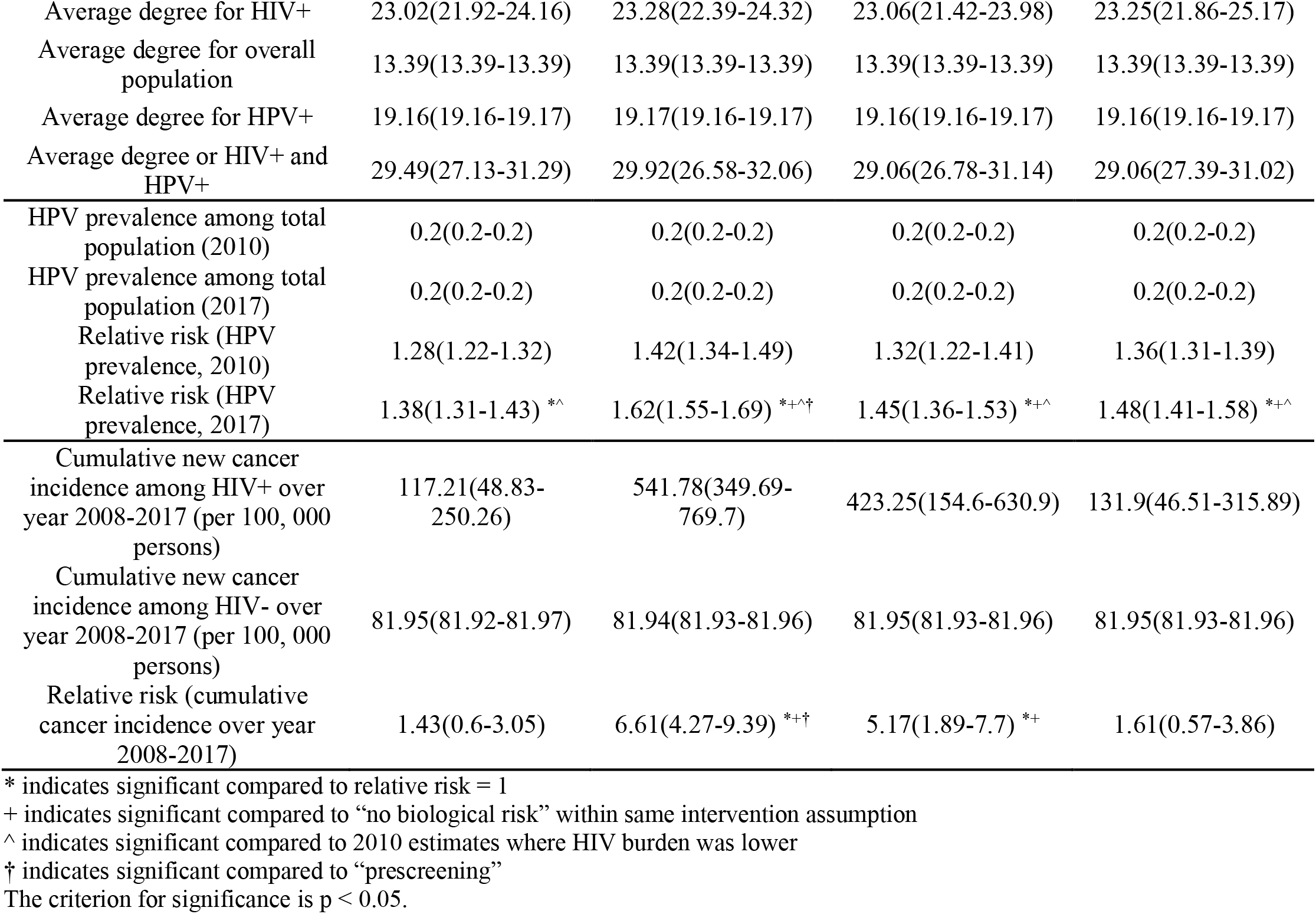
Numerical analyses of HIV and HPV disease burden and relevant network metrics under “screening only” assumption (HETF)

**Table 1c:** Numerical analyses of HIV and HPV disease burden and relevant network metrics under “status quo” assumption (HETF)

|  | Status quo |  |  |  |
| --- | --- | --- | --- | --- |
|  | No biological risk | All increased risk | Increased risk of progression | Increased risk of HPV acquisition |
| Average degree for HIV+ | 23.45(22.75-23.59) | 23.56(22.89-23.85) | 23.14(22.6-24.71) | 23.2(22.17-24.14) |
| Average degree for overall population | 13.39(13.39-13.39) | 13.39(13.39-13.39) | 13.39(13.39-13.39) | 13.39(13.39-13.39) |
| Average degree for HPV+ | 19.22(19.21-19.23) | 19.22(19.22-19.23) | 19.22(19.21-19.23) | 19.22(19.21-19.22) |
| Average degree or HIV+ and HPV+ | 28.9(27.72-32.4) | 29.1(28.2-31.15) | 29.48(27.36-31.29) | 28.63(26.02-30.69) |
| HPV prevalence among total population (2010) | 0.2(0.2-0.2) | 0.2(0.2-0.2) | 0.2(0.2-0.2) | 0.2(0.2-0.2) |
| HPV prevalence among total population (2017) | 0.19(0.19-0.19) | 0.19(0.19-0.19) | 0.19(0.19-0.19) | 0.19(0.19-0.19) |
| Relative risk (HPV prevalence, 2010) | 1.34(1.29-1.38) | 1.44(1.4-1.5) | 1.33(1.22-1.45) | 1.36(1.33-1.42) |
| Relative risk (HPV prevalence, 2017) | 1.41(1.36-1.44) <sup>*^</sup> | 1.67(1.64-1.71) <sup>*+^†</sup> | 1.48(1.39-1.58) <sup>*+^</sup> | 1.5(1.43-1.55) <sup>*+^</sup> |
| Cumulative new cancer incidence among HIV+ over year 2008-2017 (per 100, 000 persons) | 119.97(102.02-263) | 450.62(322.09-554.08) | 534.86(254.8-942.14) | 126.94(0-415.54) |
| Cumulative new cancer incidence among HIV- over year 2008-2017 (per 100, 000 persons) | 81.91(81.91-81.92) | 81.9(81.89-81.91) | 81.91(81.89-81.92) | 81.91(81.89-81.93) |
| Relative risk (cumulative cancer incidence over year 2008-2017) | 1.46(1.25-3.21) <sup>*</sup> | 5.5(3.93-6.77) <sup>*+†</sup> | 6.53(3.11-11.5) <sup>*+</sup> | 1.55(0-5.07) |

\* indicates significant compared to relative risk = 1
+ indicates significant compared to “no biological risk” within same intervention assumption
^ indicates significant compared to 2010 estimates where HIV burden was lower
† indicates significant compared to “prescreening”
The criterion for significance is $p < 0.05$ .

**Table 2a:**
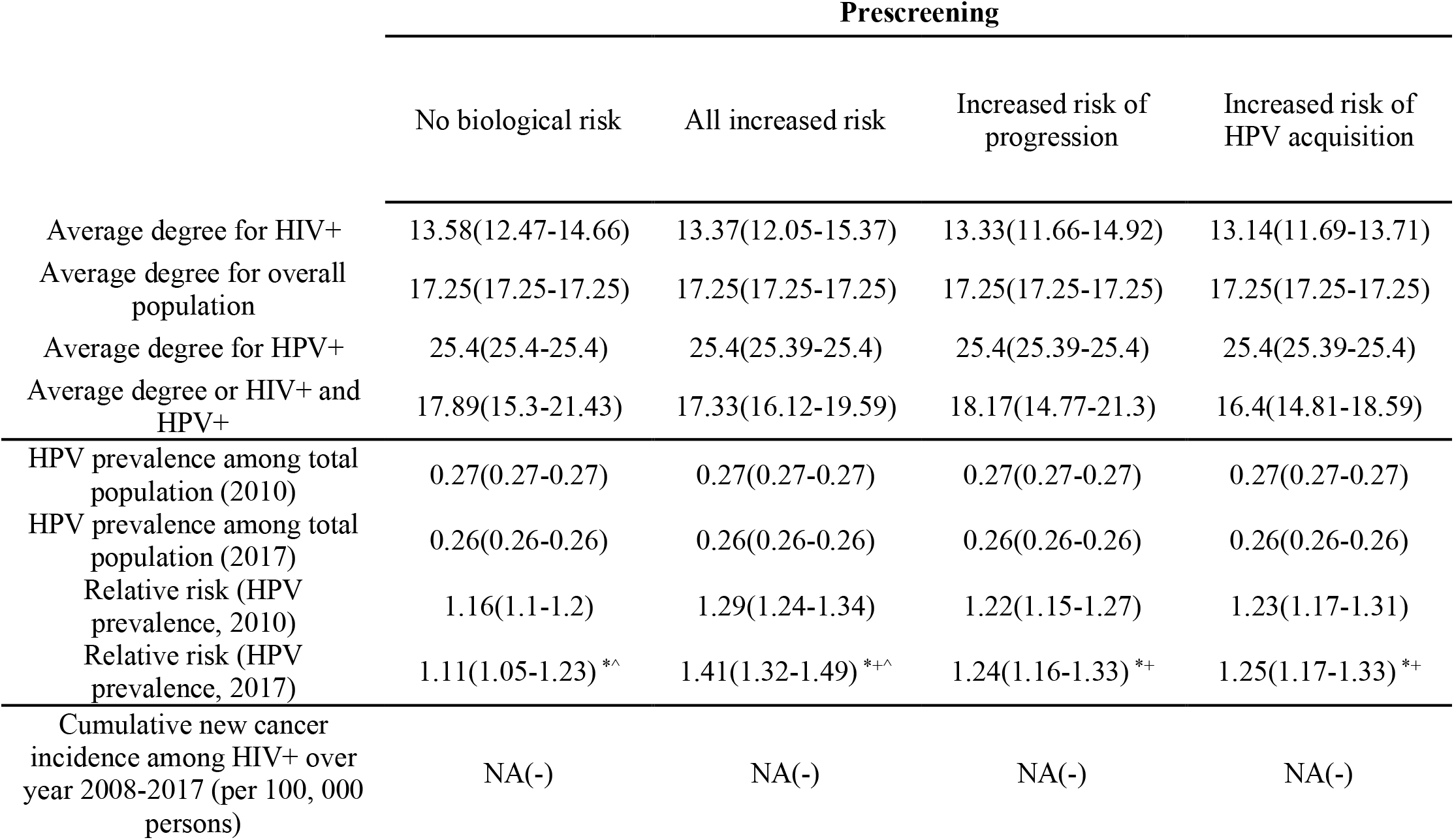
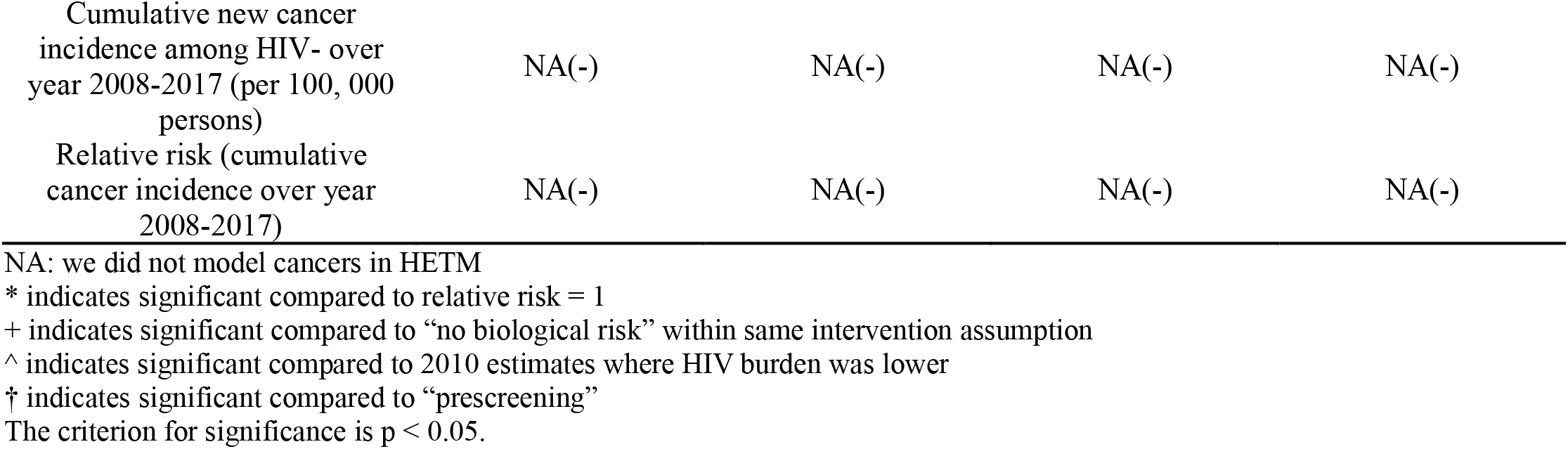
Numerical analyses of HIV and HPV disease burden and relevant network metrics under “prescreening” assumption (HETM)

|  | Prescreening |  |  |  |
| --- | --- | --- | --- | --- |
|  | No biological risk | All increased risk | Increased risk of progression | Increased risk of HPV acquisition |
| Average degree for HIV+ | 13.58(12.47-14.66) | 13.37(12.05-15.37) | 13.33(11.66-14.92) | 13.14(11.69-13.71) |
| Average degree for overall population | 17.25(17.25-17.25) | 17.25(17.25-17.25) | 17.25(17.25-17.25) | 17.25(17.25-17.25) |
| Average degree for HPV+ | 25.4(25.4-25.4) | 25.4(25.39-25.4) | 25.4(25.39-25.4) | 25.4(25.39-25.4) |
| Average degree or HIV+ and HPV+ | 17.89(15.3-21.43) | 17.33(16.12-19.59) | 18.17(14.77-21.3) | 16.4(14.81-18.59) |
| HPV prevalence among total population (2010) | 0.27(0.27-0.27) | 0.27(0.27-0.27) | 0.27(0.27-0.27) | 0.27(0.27-0.27) |
| HPV prevalence among total population (2017) | 0.26(0.26-0.26) | 0.26(0.26-0.26) | 0.26(0.26-0.26) | 0.26(0.26-0.26) |
| Relative risk (HPV prevalence, 2010) | 1.16(1.1-1.2) | 1.29(1.24-1.34) | 1.22(1.15-1.27) | 1.23(1.17-1.31) |
| Relative risk (HPV prevalence, 2017) | 1.11(1.05-1.23) <sup>*^</sup> | 1.41(1.32-1.49) <sup>*+^</sup> | 1.24(1.16-1.33) <sup>*+</sup> | 1.25(1.17-1.33) <sup>*+</sup> |
| Cumulative new cancer incidence among HIV+ over year 2008-2017 (per 100, 000 persons) | NA(-) | NA(-) | NA(-) | NA(-) |
| Cumulative new cancer incidence among HIV- over year 2008-2017 (per 100, 000 persons) | NA(-) | NA(-) | NA(-) | NA(-) |
| Relative risk (cumulative cancer incidence over year 2008-2017) | NA(-) | NA(-) | NA(-) | NA(-) |

NA: we did not model cancers in HETM
\* indicates significant compared to relative risk = 1

**Table 2b:**
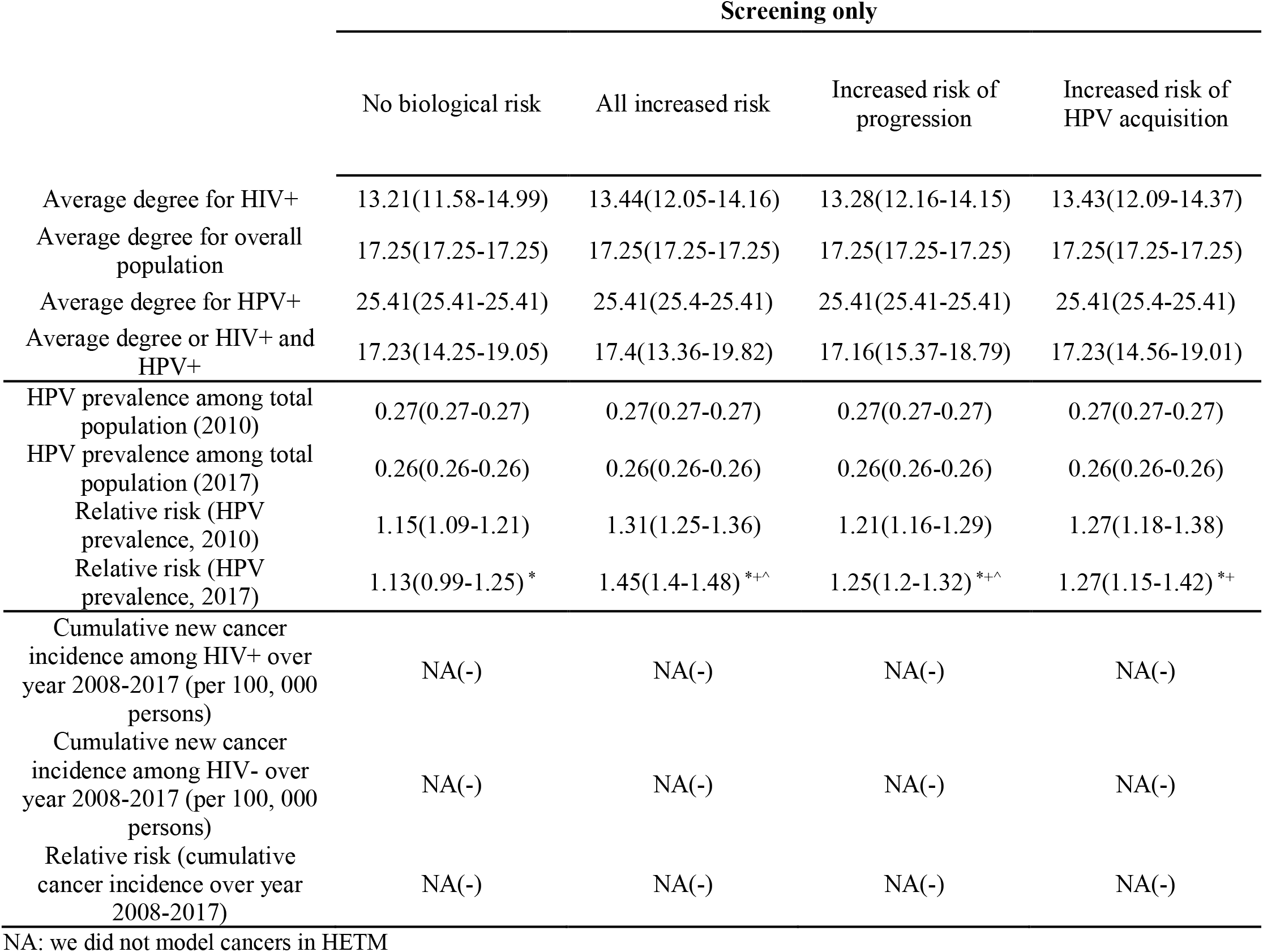
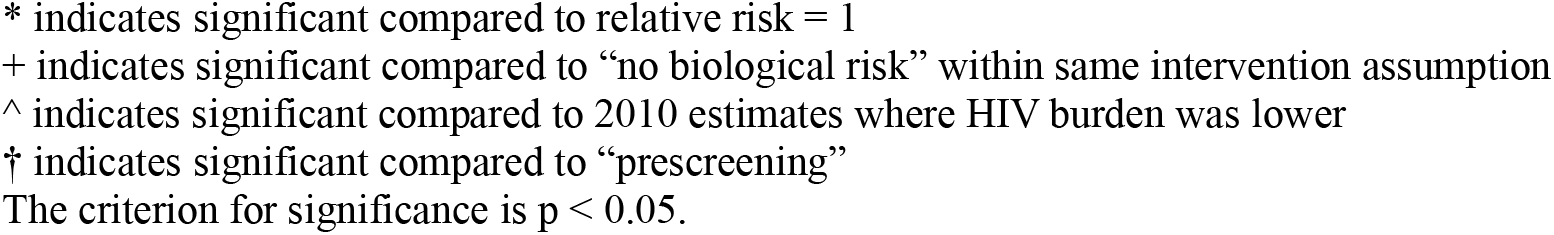
Numerical analyses of HIV and HPV disease burden and relevant network metrics under “screening only” assumption (HETM)

|  | Screening only |  |  |  |
| --- | --- | --- | --- | --- |
|  | No biological risk | All increased risk | Increased risk of progression | Increased risk of HPV acquisition |
| Average degree for HIV+ | 13.21(11.58-14.99) | 13.44(12.05-14.16) | 13.28(12.16-14.15) | 13.43(12.09-14.37) |
| Average degree for overall population | 17.25(17.25-17.25) | 17.25(17.25-17.25) | 17.25(17.25-17.25) | 17.25(17.25-17.25) |
| Average degree for HPV+ | 25.41(25.41-25.41) | 25.41(25.4-25.41) | 25.41(25.41-25.41) | 25.41(25.4-25.41) |
| Average degree or HIV+ and HPV+ | 17.23(14.25-19.05) | 17.4(13.36-19.82) | 17.16(15.37-18.79) | 17.23(14.56-19.01) |
| HPV prevalence among total population (2010) | 0.27(0.27-0.27) | 0.27(0.27-0.27) | 0.27(0.27-0.27) | 0.27(0.27-0.27) |
| HPV prevalence among total population (2017) | 0.26(0.26-0.26) | 0.26(0.26-0.26) | 0.26(0.26-0.26) | 0.26(0.26-0.26) |
| Relative risk (HPV prevalence, 2010) | 1.15(1.09-1.21) | 1.31(1.25-1.36) | 1.21(1.16-1.29) | 1.27(1.18-1.38) |
| Relative risk (HPV prevalence, 2017) | 1.13(0.99-1.25) * | 1.45(1.4-1.48) *+^ | 1.25(1.2-1.32) *+^ | 1.27(1.15-1.42) *+ |
| Cumulative new cancer incidence among HIV+ over year 2008-2017 (per 100, 000 persons) | NA(-) | NA(-) | NA(-) | NA(-) |
| Cumulative new cancer incidence among HIV- over year 2008-2017 (per 100, 000 persons) | NA(-) | NA(-) | NA(-) | NA(-) |
| Relative risk (cumulative cancer incidence over year 2008-2017) | NA(-) | NA(-) | NA(-) | NA(-) |

NA: we did not model cancers in HETM
\* indicates significant compared to relative risk = 1 + indicates significant compared to “no biological risk” within same intervention assumption ^ indicates significant compared to 2010 estimates where HIV burden was lower † indicates significant compared to “prescreening” The criterion for significance is $p < 0.05$ .

**Table 2c:** Numerical analyses of HIV and HPV disease burden and relevant network metrics under “status quo” assumption (HETM)

|  | Status quo |  |  |  |
| --- | --- | --- | --- | --- |
|  | No biological risk | All increased risk | Increased risk of progression | Increased risk of HPV acquisition |
| Average degree for HIV+ | 13.03(12.62-14.59) | 12.85(11.93-14.81) | 13.4(11.33-15.1) | 13.48(12.35-15.17) |
| Average degree for overall population | 17.25(17.25-17.25) | 17.25(17.25-17.25) | 17.25(17.25-17.25) | 17.25(17.25-17.25) |
| Average degree for HPV+ | 25.46(25.46-25.46) | 25.45(25.45-25.46) | 25.46(25.45-25.46) | 25.46(25.45-25.46) |
| Average degree or HIV+ and HPV+ | 17.53(15.75-21.03) | 16.66(14.7-20.28) | 17.67(13.36-20.53) | 17.47(15.41-19.72) |
| HPV prevalence among total population (2010) | 0.27(0.27-0.27) | 0.27(0.27-0.27) | 0.27(0.27-0.27) | 0.27(0.27-0.27) |
| HPV prevalence among total population (2017) | 0.26(0.26-0.26) | 0.26(0.26-0.26) | 0.26(0.26-0.26) | 0.26(0.26-0.26) |
| Relative risk (HPV prevalence, 2010) | 1.17(1.14-1.21) | 1.36(1.32-1.38) | 1.22(1.19-1.28) | 1.26(1.2-1.31) |
| Relative risk (HPV prevalence, 2017) | 1.13(1.01-1.16) *^ | 1.46(1.41-1.54) *+^ | 1.25(1.13-1.34) *+ | 1.32(1.22-1.39) *+^† |
| Cumulative new cancer incidence among HIV+ over year 2008-2017 (per 100, 000 persons) | NA(-) | NA(-) | NA(-) | NA(-) |
| Cumulative new cancer incidence among HIV- over year 2008-2017 (per 100, 000 persons) | NA(-) | NA(-) | NA(-) | NA(-) |
| Relative risk (cumulative cancer incidence over year 2008-2017) | NA(-) | NA(-) | NA(-) | NA(-) |

NA: we did not model cancers in HETM
\* indicates significant compared to relative risk = 1 + indicates significant compared to “no biological risk” within same intervention assumption ^ indicates significant compared to 2010 estimates where HIV burden was lower † indicates significant compared to “prescreening” The criterion for significance is $p < 0.05$ .

**Table 3a:** Numerical analyses of HIV and HPV disease burden and relevant network metrics under “prescreening” assumption (MSM)

|  | Prescreening |  |  |  |
| --- | --- | --- | --- | --- |
|  | No biological risk | All increased risk | Increased risk of progression | Increased risk of HPV acquisition |
| Average degree for HIV+ | 25.57(24.93-26.38) | 25.61(24.81-26.54) | 25.61(25.03-26.24) | 25.56(24.57-26.29) |
| Average degree for overall population | 19.7(19.63-19.82) | 19.71(19.6-19.84) | 19.7(19.64-19.79) | 19.71(19.65-19.81) |
| Average degree for HPV+ | 28.67(28.54-28.8) | 28.84(28.64-29.08) | 28.86(28.58-29.11) | 28.65(28.44-28.85) |
| Average degree or HIV+ and HPV+ | 34.67(33.2-35.35) | 34.88(33.11-35.59) | 35.63(33.77-37.22) | 33.88(32.65-34.94) |
| HPV prevalence among total population (2010) | 0.28(0.28-0.28) | 0.29(0.29-0.29) | 0.28(0.28-0.28) | 0.28(0.28-0.28) |
| HPV prevalence among total population (2017) | 0.27(0.27-0.27) | 0.29(0.28-0.29) | 0.27(0.27-0.27) | 0.28(0.28-0.28) |
| Relative risk (HPV prevalence, 2010) | 1.25(1.23-1.29) | 1.43(1.41-1.44) | 1.28(1.26-1.32) | 1.36(1.33-1.38) |
| Relative risk (HPV prevalence, 2017) | 1.33(1.31-1.37) *<sup>^</sup> | 1.62(1.58-1.66) *<sup>+^</sup> | 1.41(1.39-1.44) *<sup>+^</sup> | 1.51(1.47-1.54) *<sup>+^</sup> |
| Cumulative new cancer incidence among HIV+ over year 2008-2017 (per 100, 000 persons) | 750.74(605.91-906.12) | 2086.12(1833.14-2418.83) | 2166.61(1755.57-2342.04) | 829.67(618.48-948.46) |
| Cumulative new cancer incidence among HIV- over year 2008-2017 (per 100, 000 persons) | 530.04(528.17-531.6) | 529.72(528.25-531.62) | 530.49(528.08-532.14) | 530.21(528.64-531.88) |
| Relative risk (cumulative cancer incidence over year 2008-2017) | 1.42(1.14-1.72) * | 3.94(3.47-4.57) *<sup>+</sup> | 4.08(3.31-4.41) *<sup>+</sup> | 1.56(1.16-1.79) * |

\* indicates significant compared to relative risk = 1
+ indicates significant compared to “no biological risk” within same intervention assumption
<sup>^</sup> indicates significant compared to 2010 estimates where HIV burden was lower
<sup>†</sup> indicates significant compared to “prescreening”
The criterion for significance is $p < 0.05$ .

**Table 3b:**
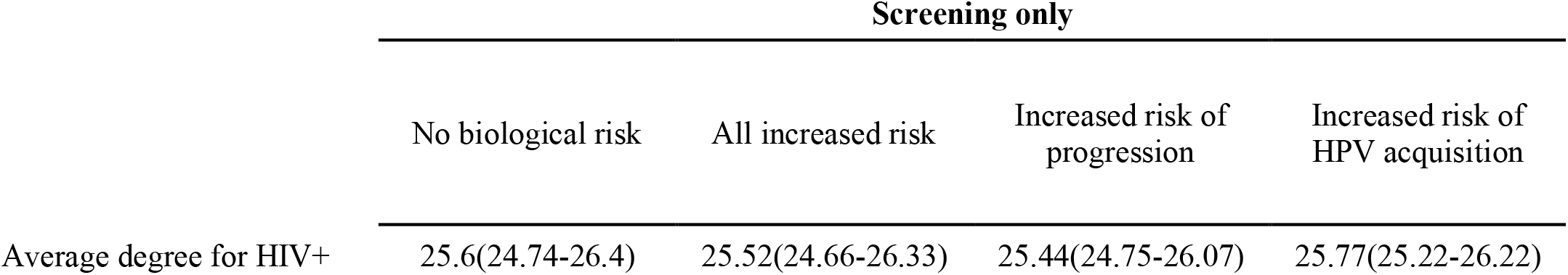
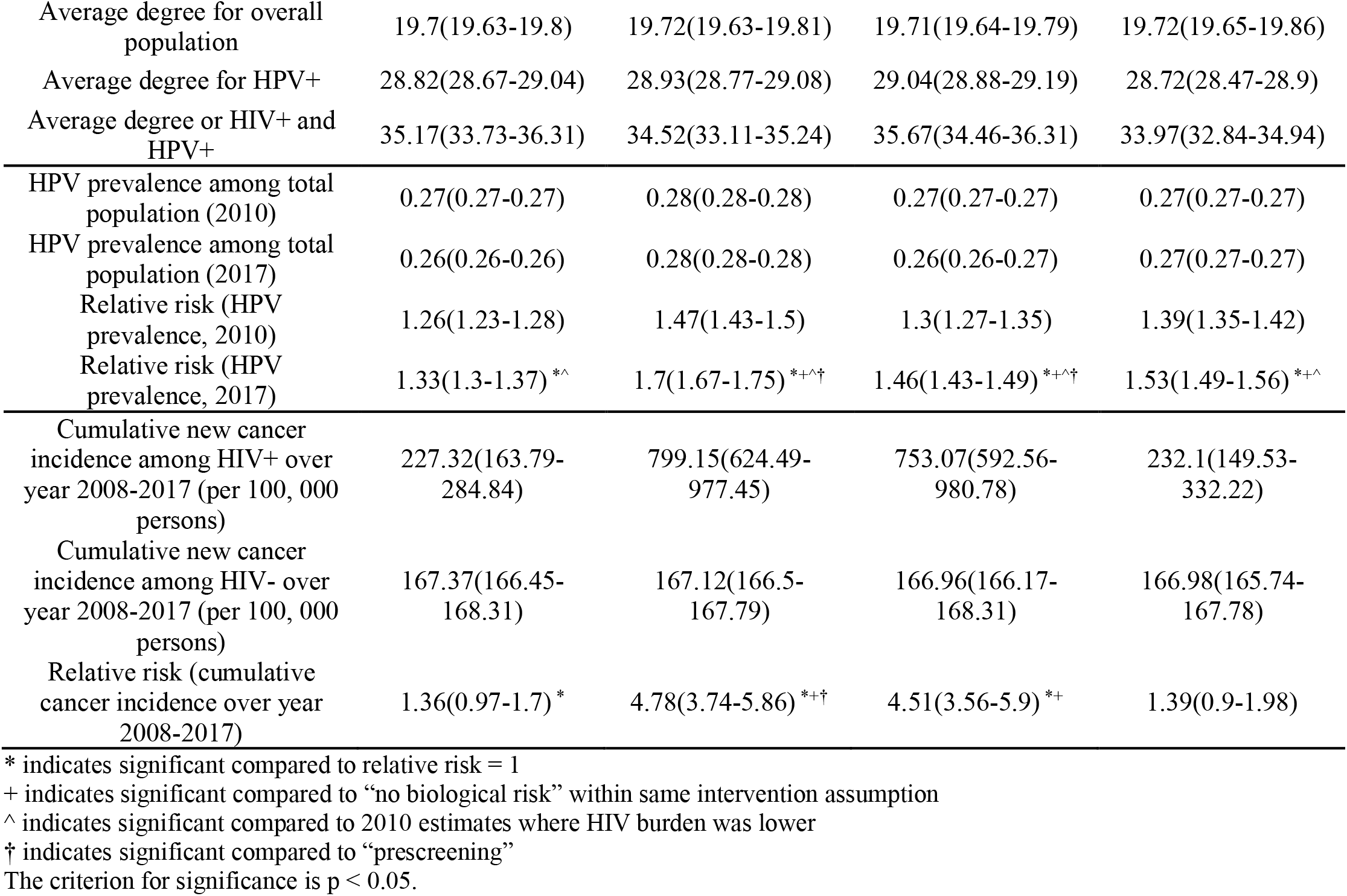
Numerical analyses of HIV and HPV disease burden and relevant network metrics under “screening only” assumption (MSM)

**Table 3c:**
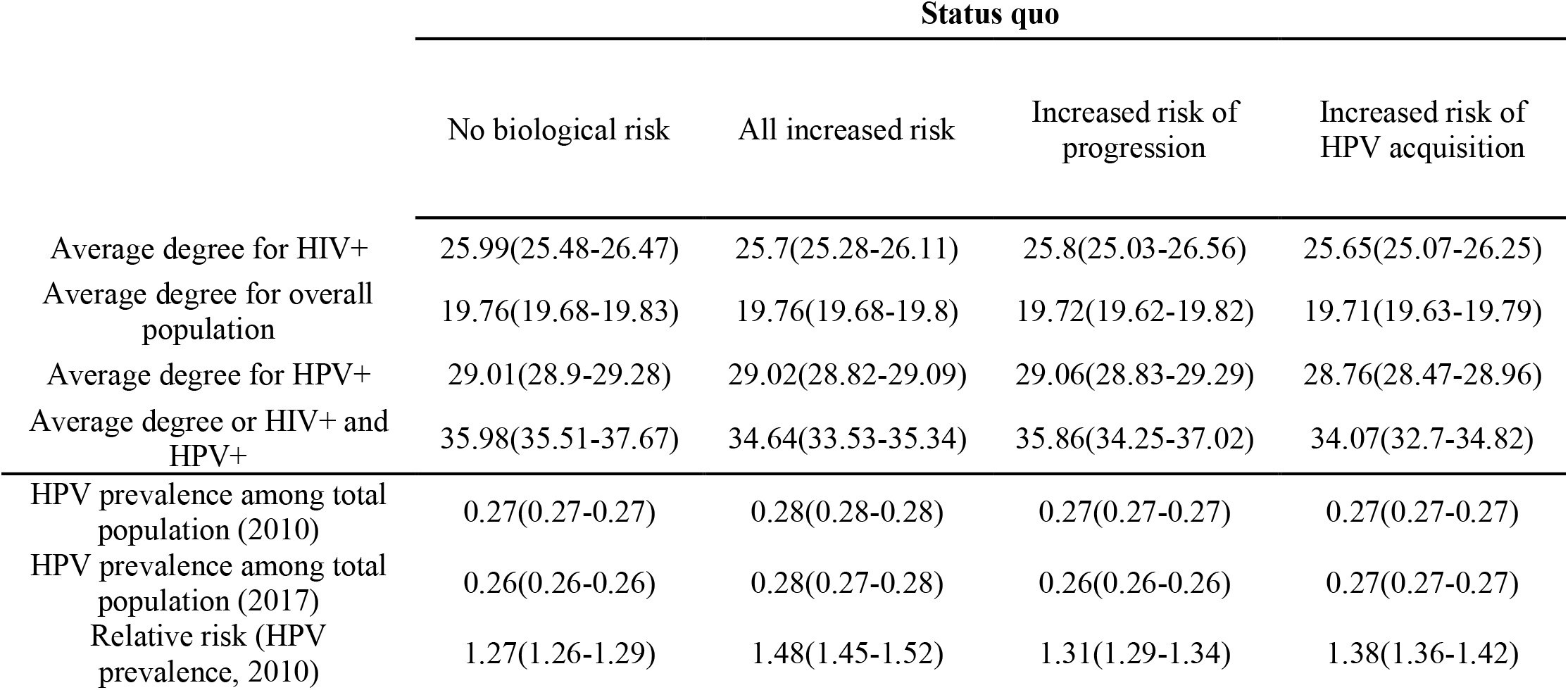
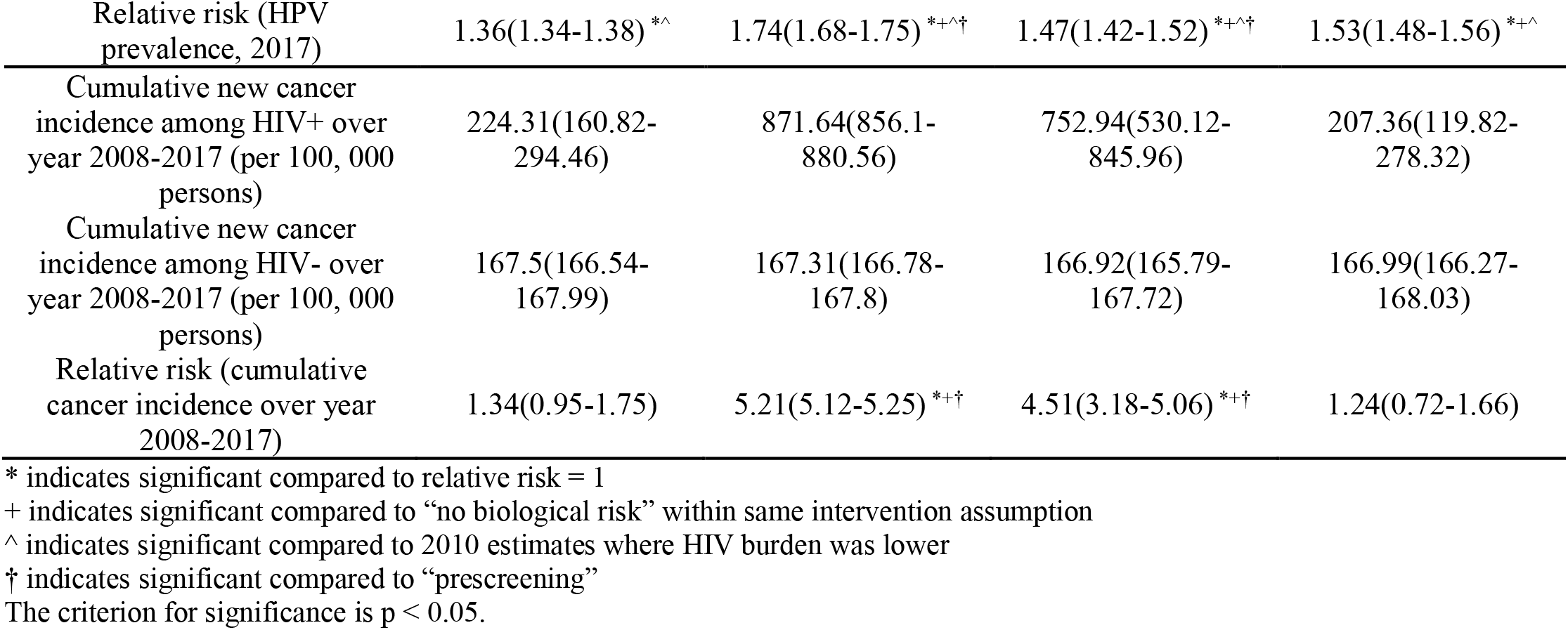
Numerical analyses of HIV and HPV disease burden and relevant network metrics under “status quo” assumption (MSM)

|  | Status quo |  |  |  |
| --- | --- | --- | --- | --- |
|  | No biological risk | All increased risk | Increased risk of progression | Increased risk of HPV acquisition |
| Average degree for HIV+ | 25.99(25.48-26.47) | 25.7(25.28-26.11) | 25.8(25.03-26.56) | 25.65(25.07-26.25) |
| Average degree for overall population | 19.76(19.68-19.83) | 19.76(19.68-19.8) | 19.72(19.62-19.82) | 19.71(19.63-19.79) |
| Average degree for HPV+ | 29.01(28.9-29.28) | 29.02(28.82-29.09) | 29.06(28.83-29.29) | 28.76(28.47-28.96) |
| Average degree or HIV+ and HPV+ | 35.98(35.51-37.67) | 34.64(33.53-35.34) | 35.86(34.25-37.02) | 34.07(32.7-34.82) |
| HPV prevalence among total population (2010) | 0.27(0.27-0.27) | 0.28(0.28-0.28) | 0.27(0.27-0.27) | 0.27(0.27-0.27) |
| HPV prevalence among total population (2017) | 0.26(0.26-0.26) | 0.28(0.27-0.28) | 0.26(0.26-0.26) | 0.27(0.27-0.27) |
| Relative risk (HPV prevalence, 2010) | 1.27(1.26-1.29) | 1.48(1.45-1.52) | 1.31(1.29-1.34) | 1.38(1.36-1.42) |
| Relative risk (HPV prevalence, 2017) | 1.36(1.34-1.38) <sup>*^</sup> | 1.74(1.68-1.75) <sup>*+^†</sup> | 1.47(1.42-1.52) <sup>*+^†</sup> | 1.53(1.48-1.56) <sup>*+^</sup> |
| Cumulative new cancer incidence among HIV+ over year 2008-2017 (per 100, 000 persons) | 224.31(160.82-294.46) | 871.64(856.1-880.56) | 752.94(530.12-845.96) | 207.36(119.82-278.32) |
| Cumulative new cancer incidence among HIV- over year 2008-2017 (per 100, 000 persons) | 167.5(166.54-167.99) | 167.31(166.78-167.8) | 166.92(165.79-167.72) | 166.99(166.27-168.03) |
| Relative risk (cumulative cancer incidence over year 2008-2017) | 1.34(0.95-1.75) | 5.21(5.12-5.25) <sup>*+†</sup> | 4.51(3.18-5.06) <sup>*+†</sup> | 1.24(0.72-1.66) |

\* indicates significant compared to relative risk = 1
+ indicates significant compared to “no biological risk” within same intervention assumption
^ indicates significant compared to 2010 estimates where HIV burden was lower
† indicates significant compared to “prescreening”
The criterion for significance is $p < 0.05$ .

The differences in *RP*_*HPV*_ could be explained through differences in the average degree in persons with HIV (*d*_*HIV*+_), average degree in persons with HPV (*d*_*HPV*+_), and average degree in the total population (*d*_*overall*_) (Table 1c, 2c, 3c). For HETF and MSM, *d*_*HIV*+_(HETF: 23, MSM: 25) and *d*_*HPV*+_(HETF: 19, MSM: 29) were significantly higher than *d*_*overall*_ (HETF: 13, MSM: 20). However, for HETM, *d*_*HPV*+_(25) was higher than *d*_*overall*_ (17) but *d*_*HIV*+_(13) was lower than *d*_*overall*_ (17). These results, as expected, suggest that if both HIV and HPV infections are concentrated in higher degree networks, then the *RP*_*HPV*_ is greater than 1. For HETM, although HPV was concentrated in higher degree networks, as the prevalence of HIV is very low, it was more randomly spread in the network, thus the *RP*_*HPV*_ was closer to 1. For HETF, compared to MSM, as HIV prevalence was moderately low, infections were focused on higher degree networks, *d*_*HIV*+_/*d*_*overall*_ was 1.77 for HETF compared to 1.25 for MSM. These results point to the sensitivity of *RR*_*HPV*_ to disease burden of both diseases and the role of network dynamics.

For all three transmission-groups and across all intervention scenarios, compared to ‘no biological risk’, *RP*_*HPV*_ was significantly higher in each of the three biological risk assumptions, lowest in ‘increased risk of HPV acquisition’ scenario, and highest increase in ‘all increased risk’ (increased risk of progression and HPV acquisition) scenario. For example, for ‘status quo’ intervention, the *RP*_*HPV*_ in ‘all increased risk’ was 1.67 (1.64-1.71) for HETF, 1.46 (1.41-1.54) for HETM, and 1.74 (1.68-1.75) for MSM (Table 1c, 2c and 3c, respectively). Whereas, for the same ‘status quo’ intervention, the *RP*_*HPV*_ in ‘no biological risk’ was 1.41(1.36-1.44) for HETF, 1.13(1.01-1.16) for HETM, and 1.36(1.34-1.38) for MSM. While the values under ‘no biological risk’ are attributable to behavioral factors, the differences between ‘all increased risk’ and ‘no biological risk’ are attributable to biological factors of coinfection.

The values of *RP*_*HPV*_ were sensitive to HIV prevalence, as seen by higher values in year 2017 compared to year 2010 for both ‘no biological risk’ and ‘all increased risk’. To note here that though HIV prevalence increased over this period, HIV care also increased (e.g., proportion on treatment with viral suppression increased from 39% in 2010 to 56% in 2017), and thus, new HIV cases were further concentrated in higher degree networks. Thus, the values of *RR*_*HPV*_ in 2017 were higher than in 2010. Further, while the fraction of increased HPV prevalence attributed to biological factors were 22%, 52%, and 43% for HETF, HETM, and MSM, respectively, in 2010, they increased to 38%, 71%, and 51% in 2017. This is as expected because the dynamics of network can amplify the overall disease risk. Thus, we can expect the fraction attributed to biological risk to also be sensitive to the network dynamics.

Relative incidence of cervical cancer (*RI*_*cancer*_) in ‘status-quo’ intervention scenario was greater than 1, and it was significantly higher in ‘all increased risk’ (3.6(1.31-6.17)) compared to ‘no biological risk’ (1.51(0.72-2.81)) (Table 1c). That is, under the assumptions used in this study, observed cases of cervical cancer incidence among women with HIV would be about 3.6 times higher than among women without HIV, about 80% of the burden attributed to biological factors and the remaining attributable to behavioral factors. Under the ‘all increased risk’ scenario, values of *RI*_*cancer*_ were slightly higher under ‘status-quo’ (3.6(1.31-6.17)) (Table 1c) compared to ‘prescreening’ (2.86(1.77-4.8)) (Table 1a), suggesting that, if screening were kept similar between women with and without HIV, the cervical cancer burden gap between them would widen. To note here, due to lack of data, we assumed screening and vaccine uptakes to be similar among persons with HIV and without HIV, although this may not be the case.

Similar results were observed for cancers among MSM (Table 3a - 3c). *RI*_*cancer*_ in ‘all increased risk’ was significantly greater than 1 in each of the three intervention scenarios, 2.83(2.33-3.51) in ‘prescreening’, 3.89(2.91-4.89) in ‘screening only’, and 4.03(3.49-4.31) in ‘status-quo’. *RI*_*cancer*_ in ‘all increased risk’ were also significantly greater than ‘no biological risk’ within each of the three intervention scenarios. To recollect that to model anal cancer among MSM, we utilized the same disease progression model as cervical cancer, for purposes of sensitivity analyses on relative risk metrics. Under this context, collectively, our results suggest higher risk of cancer among persons with HIV compared to persons without HIV, most attributed to biological risk [11–13], and a smaller but significant fraction attributed to behavioral factors.

## Discussion and Conclusions

We demonstrate the feasibility of application of a new MAC framework for jointly simulating diseases of varying prevalence but with common modes of transmission. Using the same network and sexual behavior, modeled at the individual-level for HIV and aggregated-level for HPV, the model was able to replicate both HIV and HPV in the U.S. population, and thus serves as proof-of-concept of the MAC simulation technique.

Estimates of *RP*_*HPV*_ in the literature are sparse, and not available for the U.S. population in recent times. *RP*_*HPV*_ estimates from our model, of 1.67 (1.64-1.71) among women in ‘all increased risk, status quo’ scenario, is similar to that in an observational study in the literature on an Italian population (1.71) [16], and lower than that in an observational study on a South African population (1.9) [14]. To note here that HPV prevalence and HIV prevalence are higher in South Africa than in the U.S. and European populations. A national study of HIV infection among adolescent girls in the U.S. conducted in 1996-97 [15], reported a *RP*_*HPV*_ of 3.3 (1.6-6.7). *RP*_*HPV*_ estimates from our model among adolescent girls (13-18 yrs) in the ‘all increased risk, prescreening’ scenario is 2.89 (1.41-6.1), and thus compares well with the national study.

Our model estimates for *RI*_*cancer*_ in ‘all increased risk, status quo’ scenario (3.6(1.31-6.17)), is similar to that in large population-based observational studies conducted on the U.S. population, 3.80 (3.48-4.15) in [19] and 4.1(2.3-6.6) in [29]. A recent global systematic review of cervical cancer reported a higher value of 5.34 (3.80-7.51) among high-income countries [18], suggesting likely differences across populations and study settings. Similarity in our model results with observational studies provides general model validity. To note here that the results are influenced by the assumptions used for modeling biological risk (Appendix Table S2), that were based on estimates in the literature [47,77,78]. Above results suggest that the literature estimates are a good fit, however, considering the sensitivity of the relative risk metrics and the fractions attributed to biological risk to disease burdens, closer calibrations specific to the population and timeline of interest can be conducted during implementation of the model for decision analyses.

Relative prevalence of HPV was greater than 1 in ‘no biological risk’ and ‘all increased risk’, values in ‘all increased risk’ being higher than ‘no biological risk’. Further, in ‘all increased risk, status-quo’ scenario, relative incidence of cervical cancer was much higher than HPV. Similar patterns were observed for cancers and HPV among MSM. These results suggest that behavioral factors contribute to increased risk of HIV-HPV coinfection and are further exacerbated by biological factors, especially for cancer cases. These results suggest the need for both behavioral interventions to reduce the risk for infection, and care interventions for early detection and treatment of HPV to reduce the risk of cancers. Social factors are among key drivers of increased risky behavior e.g., higher number of partners and higher condomless sex, or lower adherence to treatment that prevents transmission by suppressing viral load [36–39]. Thus, structural interventions such as healthcare coverage, subsidized housing and food programs, and access to mental healthcare [42–45,83], are key part of behavioral interventions.

Model estimates of relative prevalence and fraction attributable to each factor (biological and behavioral) varied with HIV burden and care, suggesting influence of network dynamics, as verified by the average degree estimates. Model estimates of relative incidence of cervical cancer was higher in the HPV/ cancer screening scenario compared to the pre-screening scenario, suggesting sensitivity to HPV burden and care. Thus, jointly modeling diseases in a dynamic network model can help more accurately measure the impact of interventions. Above results also support the need for more focused screening among persons with HIV (Note here that, to evaluate the sensitivity to changes in interventions, our model assumed same levels of screening among persons with and without HIV).

Our work is subject to limitations. We only focused on a general model-fit for HPV among women and did not conduct detailed sensitivity analyses for the robustness of the calibration method. Though the overall HPV model provided a good fit in most metrics in the pre-screening results (Fig 2 and Fig 3a), the results under the screening scenario (Fig 3b) were not always same as surveillance estimates. As post-screening model fits are largely influenced by assumptions for age-specific compliance to screening, and not disease epidemiology, considering the scope of our analyses we did not attempt to calibrate screening rates. Further, due to unavailability of more recent data on screening rates, we kept it constant at the 2006 data assumptions for years 2006 to 2017. These assumptions are acceptable because, as noted earlier, per surveillance data, there were minor changes in cervical cancer incidence and mortality over the period 2006 to 2017. We did not attempt to calibrate biological risk multipliers but used estimates from the literature. Though the model outcomes of relative risk were in the range of that reported from observational studies, considering its sensitivity to disease burden and care changes over time, during implementation of model to inform decisions it should be specifically calibrated to the population under study. For MSM, we did not specifically model anal cancer (the sequelae of most concern) but assumed similar epidemiology to cervical cancer. As observations here justify the need for jointly modeling diseases, as opposed to independent disease models, adding anal cancer in future work would be a suitable extension. We only evaluated the increased risk of HPV among persons with HIV and did not evaluate vice-versa.

We believe the overall validation of HIV and HPV serves as proof-of-concept of the MAC framework for joint modeling related diseases with widely varying prevalence but spread on a common network. The model can be expanded to include other STIs. As numerical analyses suggest, disease interactions are attributed to both behavioral factors and biological factors. Structural interventions that address social determinants are a key part of behavioral interventions. The MAC framework is suitable for modeling behaviors as a function of social determinants, and further, measuring the impact of structural interventions on prevention of overall STI burden. The network features of the MAC framework are suitable for network-based analyses of interventions such as cluster detection and response, that has been successful in identifying populations with high HIV transmission [59,84]. Expanding HIV cluster-based network detection to joint-disease framework could help identify populations most vulnerable to diseases, their intervention needs, and collectively evaluate the impact of intervention on overall disease prevention.

Our model is also suitable for joint modeling of sub-populations with widely varying disease burden, but who interact with each other, such as HETF, HETM, and MSM in this case study. Recent studies highlight the high burden of HPV-related clinical conditions among HIV infected MSM [2,85,86], but there are only few models in this area [87]. Further, they model MSM in isolation. The computational tractability of the MAC simulation technique makes joint modeling sub-populations with widely vary prevalence feasible. HIV burden is disproportionately higher among MSM in the U.S. [82], and there are significant associations of HIV infections among HETF from mixing with MSM [56,57], which is also relevant in the context of higher burden of cervical cancer among women with HIV. Thus, simulation of interactions between sub-populations and between diseases can more accurately represent the dynamics of infection spread. Behaviors leading to transmission across these sub-groups are driven by social stressors and socio-economic vulnerability and are typically exacerbated in neighborhoods of high poverty [88,89]. Therefore, the model developed here would be suitable for jointly evaluating combinations of structural and disease-specific interventions, across sub-population groups, and across multiple diseases. Simulating these in a national context could help inform broader public health policies.

## Data Availability

All data produced in the present study are available in the manuscript and supporting information

## Acknowledgments

This work was funded by the National Science Foundation under grant #1915481. The funding agreement ensured the authors’ independence in designing the study, interpreting the data, writing, and publishing the report.

