## Supplemental appendix for "Joint modeling HIV and HPV using a new hybrid agent-based network and compartmental simulation technique"

### S1 Overview of MAC

The mixed agent-based compartmental (MAC) simulation framework was developed for co-modeling of diseases spread over a common contact network but have varying levels of prevalence and incidence that neither agent-based nor compartmental model alone is sufficient. The computational framework for the hybrid agent-based compartmental simulation was enabled through use of a recently developed agent-based evolving network modeling technique (ABENM) [1], applied to the development of the Progression and Transmission of HIV (PATH 4.0) model in the U.S. and validated against data from the National HIV Surveillance Systems (NHSS) [2]. The main concept of ABENM is to simulate only persons infected with at least one lower prevalence disease and their immediate contacts at the individual level as agents of the simulation, and to model all other persons including those with higher prevalence diseases using a compartmental modeling structure. Immediate contacts are defined as all partners a person will have over their lifetime who at the current time-step may be infected or susceptible. As these contacts in the network become infected with a lower prevalence disease, their immediate contacts are added as agents to the network (transitioning from the compartmental portion of the model to the network portion of the model), thus *evolving the contact network*. An Evolving Contact Network Algorithm (ECNA) maintains the network dynamics. Here we provide an overview of the MAC simulation framework, without loss of generality, using a two-disease example, lower prevalence Disease 1 and higher prevalence Disease 2. In the analyses presented in the main paper, we modeled HIV as Disease 1, adopting the validated PATH 4.0 model [2] and HPV. We believe this framework can be generalized to any number of diseases, the computational complexity and relevance informing decisions for modeling it in the ABENM or in the compartmental.

#### *S1.1 Overview of MAC using lower prevalence disease 1 and higher prevalence disease 2*

We present the framework using HIV as Disease 1 and a hypothetical Disease 2 for simplicity. More details on modeling HPV can be found in Section 2.2. We track HIV-infected persons and immediate contacts using a dynamic graph $G_{t}\mathcal{(N,E)}$, with the number of nodes in the graph $Q_{t}=\left| \mathcal{N(}G_{t}) \right|$ and the number of edges $\mathcal{|E}\left( G_{t} \right)|$ dynamically changing over time $t$ as persons become newly infected with HIV and their immediate contacts are added to the network. Each person in the network has attributes: age, transmission-group (heterosexual female (HETF), heterosexual male (HETM), men who have sex with men (men only and men and women) (MSM), degree (number of lifetime partnerships), geographic jurisdiction, health status (Disease 1 no Disease 2, Disease 1 Disease 2, no Disease 1 no Disease 2), and HIV-related disease and care continuum stages.

We track all other persons using an array $S_{t}$ of size $A\times R\times D\times\mathcal{G}X H$.

Where,

$A$ is the number of age-groups,

$R$ is the number of transmission risk groups,

$D$ is the number of degree-bins (degree is the number of contacts per person, degrees are grouped into bins analogous to age grouped into age-groups),

$\mathcal{G}$ is the number of geographic jurisdictions (equal to 1 here corresponding to national), and

$H$ is the number of health states (two in numerical analyses: no Disease 1 no Disease 2, and Disease 1 no Disease 2).

Therefore, each element of the array ($S_{t}\left[ \bar{a}, r, \bar{d},g,h \right])$ is the number of people in that specific category, and thus, $Q_{t}+\sum_{i\in\bar{a}, r, \bar{d},g,h} S_{t,i}$, would be the total number of people in the population at time $t.$

As all HIV infected persons and exposed partners are in the network, HIV transmissions and HIV disease progression are modeled at the individual level (using HIV transmission and HIV disease progression modules in Section 1.3). Disease 2 transmission and progression are modeled using differential equations (as typically done in compartmental modeling technique) but with the consideration that people in both the network $G_{t}\mathcal{(N,E)}$ and compartmental array $S_{t}$ can be infected with Disease 2 (see compartmental module in Section 1.3).

The mathematical challenges to address for this method to work is to maintain the dynamics between the network $G_{t}\mathcal{(N,E)}$ and compartmental array $S_{t}$, including, transitioning people from $S_{t}$ to $G_{t}\mathcal{(N,E)}$. Specifically, determining ‘who’, i.e., the degree, transmission risk group, age, and geographical location of the persons, are to be added as the immediate contacts of the node newly infected with HIV, as these characteristics of infected persons and their contacts are known to be correlated [3]. Upon determining and adding all lifetime partners of nodes newly infected with HIV, we need to determine when the partnerships would initiate and dissolve, including the age of both partners at that time, such that the overall dynamics of age-mixing and risk-group-mixing of the resulting network match that of the U.S. population over time. As network generation techniques in commonly used agent-based models are designed to generate the network between the full population (susceptible and infected persons), they cannot be adopted here. We developed an evolving contact network algorithm (ECNA) for modeling the transitions from $S_{t}$ to $G_{t}\mathcal{(N,E)}$ (see ECNA module in Section 1.3.3). As HIV is a chronic infection, once persons enter the network $G_{t}\mathcal{(N,E)}$ they do not transition back to $S_{t}$. However, modeling such a transition (for cases of lower prevalence disease that are not chronic) would be simple as they can be added to the compartments corresponding to their group.

We discuss the computational structure of MAC in Section 1.2 and the four simulation modules in Section 1.3. A visual representation of the computational structure is presented in Figure 1 (main manuscript). All notations used in the model are also summarized in Table S1.

#### *S1.2 Computational structure of MAC*

As noted above, following the compartmental modeling structure, we use a five-dimensional array $S_{t}$ to keep track of the number of susceptible persons (who are not contacts of persons with HIV infection), i.e., $S_{t}[\bar{a}, r, \bar{d},g,h]$ is the number of susceptible persons in age-group $\bar{a}$ , transmission-group $r$, degree-bin $\bar{d}$, pseudo-geographic jurisdiction $g$, and health status $h$at time $t$.

As noted above, following the ABNEM structure, we use a dynamic graph $G_{t}\mathcal{(N,E)}$ to track HIV-infected persons and their immediate contacts, where $\mathcal{N}$ is a set of nodes, each node representing an HIV-infected person or a susceptible sexual partner (they may or may not have Disease 2), and$\mathcal{E(}G_{t})$ is a set of undirected edges, an edge $\left\{ i,j \right\}$ representing a sexual partnership between nodes $i$ and $j$.

The graph $G_{t}\mathcal{(N,E)}$ has the following features:

**Static adjacency matrix**: $C_{t}$ of time-variant size $Q_{t}\times Q_{t},$ with static elements $C_{t}[i,j]=1$ if $i$ and $j$ are sexual partners anytime during their lifetime and $C_{t}[i,j]=$0 otherwise, and

**Dynamic adjacency matrix**: $V_{t}$ of time-variant size $Q_{t}\times Q_{t}$, with element $V_{t}[i,j]=1$ if $i$ and $j$ are in a partnership during month $t$ and $V_{t}[i,j]=0$ otherwise.

Each edge $\left\{ i,j \right\}\epsilon\mathcal{E}$has the following features (similar to nodes having features of say age, sex, etc., edges can also have features):

**Partnership initiation time**:$\overline{t}(\left\{ i,j \right\})$ representing the simulation month for when the partnership initiated,

**Partnership termination time:** $\underline{t}(\left\{ i,j \right\})$ representing the simulation month when the partnership terminated,

**Partnership initiation** **age:** $\left\{ {\bar{\mathcal{a}}}_{i},{\bar{\mathcal{a}}}_{j} \right\}$ representing the age of nodes $i \mathrm{and}j$, at the time of partnership initiation, and

**Partnership termination** **age:** $\left\{ {\underline{\mathcal{a}}}_{i},{\underline{\mathcal{a}}}_{j} \right\}$ representing the age of nodes $i \mathrm{and}j$, at the time of partnership termination.

Each node $j\epsilon\mathcal{N}\left( G_{t} \right)$ has the following features:

**Actual degree**:$d_{j}$ representing the actual number of lifetime sexual partners of node $j$,

**Current degree:** $\hat{d}_{t,j}$ representing the number of lifetime sexual partners of person $j$ who are already added as nodes in $G_{t}\mathcal{(N,E)}$; if node $j$ is HIV-infected $\hat{d}_{t,j}=d_{j}$, if node $j$ is HIV-susceptible $\hat{d}_{t,j}\leq d_{j}$, and thus dynamically changing with time $t$,

**Partnership distribution matrix:** $L_{t,j}$ of size $A\times2$, where $A$ is the number of age-groups, $L_{t,j}[\overline{a},1]$ is the number of partnerships that node $j$ initiates in age group $\overline{a}$, and $L_{t,j}[\overline{a},2]$ is the number of partnerships that are yet to be assigned; the subscript $t$ is used to indicate that the values of column 2 of $L_{t,j}$ can change over time, specifically, $L_{t,j}[\overline{a},2]$ is a column of zeros if the node is HIV-infected as all their partnerships are already assigned, and greater than or equal to zero if the node is HIV-susceptible (when the HIV-susceptible person is added as a contact of a different HIV-infected person one of the rows is decremented, and when the HIV-susceptible person becomes HIV-infected all rows of column 2 are decremented to zero as their partners are found and added - the HIV-ECNA was specifically developed for determining when and how to assign these partnerships, and thus generating the network, which is discussed in Section 1.3.3.),

**HIV-infection status**: $\mathcal{h}_{t,j}=1$ if node $j$ is HIV-infected and $\mathcal{h}_{t,j}=0$ otherwise,

**Disease 2-infection status:** $\bar{h}_{t,j}=1$ if node $j$ is Disease 2-infected and $\bar{h}_{t,j}=0$ otherwise

**Deceased status**: $\mathcal{m}_{t,j}=1$ if node $j$ is alive and $0$ otherwise,

**Age**: $\mathcal{a}_{t,j}$ taking an integer value representative of the age of node $j$,

**Pseudo-geographic jurisdiction**: $g_{j}$ taking an integer value representative of the pseudo-geographic location of node $j$,

**Transmission-group**: $\mathcal{r}_{j}$ taking one of the following values, representative of risk-group of node $j$, $\mathcal{r}_{j} \in\{$heterosexual female, heterosexual male, MSM}, and

**HIV care continuum and disease stage**: $\mathcal{s}_{t,j}$ taking one of the following values, 0 (not infected), 1 (infected, acute HIV stage, and undiagnosed), 2 (non-acute HIV, and undiagnosed), 3 (diagnosed and not in care), 4 (in care not on antiretroviral therapy (ART) treatment), 5 (on ART no viral load suppression (VLS)), or 6 (on ART with VLS).

The main relationships between different components of the graph $G_{t}\mathcal{(N,E)}$ are the following.

Between partnership initiation $\overline{t}\left( \left\{ i,j \right\} \right)$ and termination $\underline{t}(\left\{ i,j \right\})$ times and static and dynamic adjacency matrices ($C_{t}$ and $V_{t}$):

$C_{t}\left[ i,j \right]=\left\{ \begin{aligned} 1 if \left\{ i,j \right\}\epsilon\mathcal{E} \\ 0 otherwise \end{aligned} \right.,$ i.e., if $\left\{ i,j \right\}$ are partners at some point during their life, this will have a value of 1,

$V_{t}[i,j]=\left\{ \begin{aligned} 1\mathrm{if} \overline{t}\left( \left\{ i,j \right\} \right)\leq t\leq\underline{t}(\left\{ i,j \right\}) \\ 0 otherwise \end{aligned} \right.$, i.e., if $\left\{ i,j \right\}$ are partners at time $t$ this will have a value of 1, and thus,

$C_{t}\left[ i,j \right]\geq V_{t}[i,j]$.

Between actual degree $d_{j}$, current degrees $\hat{d}_{t,j}$, and static adjacency matrix $C_{t}:$

$\hat{d}_{t,j}\left\{ \begin{aligned} =d_{j} if node j is infected \\ \leq d_{j} if node j is susceptible \end{aligned} \right.$ , i.e., if a node is infected, they are linked to all partners they will have (actual degree) over their lifetime and thus $d_{j}=\hat{d}_{t,j}$, and if a node is susceptible, they are only linked to their infected partners and thus $d_{j}\leq\hat{d}_{t,j}$, and

$\hat{d}_{t,j}=\sum_{i=1:Q_{t}} C_{t}\left[ i,j \right]$, i.e., $C_{t}$ keeps track of their current degree.

Between actual degree $d_{j}$ and partnership distribution matrix $L_{t,j}$:

$\sum_{\overline{a}=1:A} L_{t,j}[\overline{a},1]=d_{j}$, at any $t$, i.e., as $L_{t,j}[\overline{a},1]$ tracks number of partnerships that initiate at age-group $\overline{a}$, when summed over all $\overline{a}$ it should add to the actual degree $d_{j}$ for all nodes whether infected or susceptible, and

$\sum_{\overline{a}=1:A} L_{t,j}[\overline{a},2]=\left\{ \begin{aligned} d_{j}-\hat{d}_{t,j} if node j is susceptible \\ 0 if node j is infected \end{aligned} \right.$, i.e., as $L_{t,j}[\overline{a},2]$ tracks number of partnerships that initiate at age-group $\overline{a}$ and are yet to be generated, $\sum_{\overline{a}=1:A} L_{t,j}[\overline{a},2]$would be zero if the node is infected because all partnerships of an infected node are already connected in the network, and would be equal to the number of partners yet to be assigned if the node is susceptible. (Assigning partnerships and all other features related to the network are part of the newly developed HIV-ECNA network generation algorithm, discussed later).

This section presented the computational structure of the model, specifically the compartmental modeling structure, the network structure, and the features of the nodes and edges in the network. The next section describes the methods (modules) used in simulating these features. We will discuss HPV and cervical cancer model, calibration and validation in Section 2 and Section 3 provides an overview of the steps of the simulation.

#### *S1.3 Four main modules of MAC*

We present the overall MAC framework through four modules, that are run at every time-step (monthly) of the simulation: a compartmental module for simulating Disease 2, a Bernoulli transmission module for simulating new infections, the ECNA network generation module for generating partnership networks of new HIV-infected persons, and a disease progression module for simulating HIV-related events for HIV-infected persons.

##### S1.3.1 Compartmental module for simulating higher prevalence disease 2

This module updates the demographic features (births, aging, and deaths) and Disease 2 features (transmission and progression) of persons tracked through the array $S_{t}$, using difference equations as follows.

$S_{t}=S_{t-1}+\frac{{dS}_{t}}{dt}\Delta t$; $\frac{{dS}_{t}}{dt}=S_{t-1}G_{t}$

$\frac{{dS}_{t}}{dt}$ is the rate of change in $S_{t}$, modeled as Markov chain state transitions through non-stationary generator array $G_{t}$. The compartmental model array $S_{t}$ for HPV is of size $7\times3\times9\times1 \times41$, corresponding to size of age-group, risk-group, degree-bin, geographical jurisdiction, and HPV health state, and thus, $G_{t}$ is of size $7\times3\times9\times1 \times41\times41$. Details of the generator matrix specific to HPV is presented in Section 2. For a generalized understanding of this equation, we make a simplified assumption of a single disease stage, i.e., $\bar{h}=0$ for disease free and $\bar{h}=1$ for Disease 2 (D2). Thus, we can expand the equation as follows

$$\frac{{dS}_{t}}{dt}\left[ \bar{a}, r, \bar{d},g,h=0 \right]=-D2 new infections+D2 recovery-aging +aging in-deaths$$

$$\frac{{dS}_{t}}{dt}\left[ \bar{a}, r, \bar{d},g,h=1 \right]=D2 new infections-D2 recovery-aging +aging in-deaths$$

Each element on the right-hand side can be calculated as follows,

$D2 new infections=\theta_{\bar{a}, r, \bar{d},g}S_{t}\left[ \bar{a}, r, \bar{d},g,h=0 \right]$,

$D2 recovery=\gamma S_{t}\left[ \bar{a}, r, \bar{d},g,h=1 \right]$ ,

$aging out$ = $\left( \frac{1}{\left| \bar{a} \right|} \right)S_{t}\left[ \bar{a}, r, \bar{d},g,h=0 \right] ,$

$aging in$ = $\left( \frac{1}{\left| \bar{a}-1 \right|} \right)S_{t}\left[ \bar{a}-1, r, \bar{d},g,h=0 \right],$ and

$deaths=\delta_{\bar{a}} S_{t}\left[ \bar{a}, r, \bar{d},g,h \right]$,

where,

$\theta_{\bar{a}, r, \bar{d},g}$is the force of infection for persons in age-group $\bar{a}$, transmission risk group $\bar{r}$, degree-bin $\bar{d}$, pseudo-geographic jurisdiction $g$at time $t$,

$\gamma$ is the recovery rate (in these hypothetical analyses we assumed it is static, but could be varied as a function of $\bar{a}, r, g$, to represent epidemiological differences by demography or as a function of time $t$ to represent changes in care or treatment over time,

$\left| \bar{a} \right|$ is the age-interval of age-group $\bar{a}$, and

$\delta_{\bar{a}}$ is mortality rate as a function of age-group $\bar{a}$ (here we only modeled all-cause mortality, but it could be varied as a function of health status $\bar{h}$).

For this simple single Disease 2 demonstration example, we can write force of infection $\theta_{\bar{a}, r, \bar{d},g}$ as follows, (for clarity, we replace $\theta_{\bar{a}, r, \bar{d},g}=i$ in subscript to denote the group of reference)

$\theta_{i}=\beta_{i}c_{i}\sum_{j} M_{ij}\frac{I_{j}}{N_{j}}$,

where,

$\beta_{i}$ is the per contact transmission probability,

$c_{i}$ is the number of contacts,

$M_{ij}$ is partnership mixing of group $i$ with group $j$ (here group represents a combination of $\bar{a}, \bar{r}, \bar{d},g$),

$I_{j}$ is the number of persons infected with Disease 2 in group $j$, and

$N_{j}$ is the number of persons in group $j$.

In this MAC approach,

$I_{\bar{a}, \bar{r}, \bar{d},g}=S_{t}\left[ \bar{a}, r, \bar{d},g,h=1 \right]+|\left\{ \mathcal{N}_{\bar{a}, \bar{r}, \bar{d},g,\bar{h}=1} \right\}|$,

$N_{\bar{a}, \bar{r}, \bar{d},g}=S_{t}\left[ \bar{a}, r, \bar{d},g,h=0 \right]+S_{t}\left[ \bar{a}, r, \bar{d},g,h=1 \right]+|\left\{ \mathcal{N}_{\bar{a}, r, \bar{d},g,h=0} \right\}+ |\left\{ \mathcal{N}_{\bar{a}, r, \bar{d},g,h=1} \right\}|$

i.e., $I_{j}$ would be the sum of Disease 2 infected persons in group $j$ ($S_{t}\left[ \bar{a}, r, \bar{d},g,h=1 \right]$) in the compartmental model and the number of nodes in the network in group $j$ ($|\left\{ \mathcal{N}_{\bar{a}, r, \bar{d},g,h=1} \right\}|$), and similarly, $N_{j}$ would be the sum of persons in group $j$ in the compartmental model and number of nodes in group $j$ in the network with Disease 2 ($\bar{h}=1$) and without Disease 2 ($\bar{h}=0$).

*Simulating Disease 2 among HIV infected persons in the network* $G_{t}\mathcal{(N,E)}$

Continuing with the simple singe stage Disease 2 demonstration example, we can use the above transition rates (infection rate $\theta_{\bar{a}, \bar{r}, \bar{d},g}$ and progression rate $\gamma$) to simulate Disease 2 among nodes in the network. We first convert rates to probability, as $probability =1-exp^{-rate.1}$ (assuming sojourn times follow exponential distribution). We then use a binomial distribution to determine number of nodes to transition, specifically,

number of nodes newly infected with Disease 2 =$i_{D2}\sim$Binomial( $\left| \left\{ \mathcal{N}_{\bar{a}, r, \bar{d},g,h=0} \right\} \right|, 1-exp^{-\theta_{\bar{a}, r, \bar{d},g} .1})$

number of nodes newly recovered from Disease 2 =$r_{D2}$~Binomial( $\left| \left\{ \mathcal{N}_{\bar{a}, r, \bar{d},g,h=1} \right\} \right|, 1-exp^{-\gamma.1})$

We then randomly choose $i_{D2}$ from node-set $\left\{ \mathcal{N}_{\bar{a}, r, \bar{d},g,h=0} \right\}$ and set their Disease 2 status $\bar{h}=1$, and randomly choose $r_{D2}$ from node-set $\left\{ \mathcal{N}_{\bar{a}, r, \bar{d},g,h=1} \right\}$ and set their Disease 2 status $\bar{h}=0$. For the HPV model, such calculations would be conducted for each of the 41 disease stages.

##### S1.3.2 Transmission module for simulating new disease 1 infections in network (HIV-infection)

This module determines if a HIV-susceptible node $l$ in the network $G_{t}\mathcal{(N,E)}$ becomes infected using a Bernoulli transmission equation and individual-level sexual behaviors and transmission risk factors, each factor modeled as functions of demographic features, and HIV testing and treatment status of infected contacts. Note, as persons in the compartmental model array $S_{t}$ are not connected to an HIV-infected person, their chance of infection is zero. However, note, persons can move from $S_{t}$ to $G_{t}\mathcal{(N,E)}$ upon becoming partners of an HIV-infected person, modeled using the HIV-ECNA algorithm discussed in the next section, which would then expose them to HIV infection. Specifically, every time-step (monthly) of the simulation, this module determines if a HIV-susceptible node $l$ in graph $G_{t}\mathcal{(N,E)}$ becomes HIV-infected i.e., for nodes with HIV infection status $\mathcal{h}_{t-1,l}=0$ it estimates its updated value $\mathcal{h}_{t,l}$ as follows.

$\mathcal{h}_{t,l}=F^{-1}\left( 1-\prod_{j=1}^{Q_{t}} \left( 1-\alpha_{j}\varepsilon\right)^{s_{t,j}.c_{t,j}}\left( 1-\alpha_{j} \right)^{s_{t,j}.(1-c_{t,j})} \right)$,

where,

$\alpha_{j}=V_{t}\left[ l,j \right]. \mathcal{h}_{t,j} .\mathcal{m}_{t,j} .p_{t,j}$, where $V_{t}\left[ l,j \right], \mathcal{h}_{t,j} ,\mathcal{m}_{t,j}$ are the elements of the graph described in Section 1.2, and $p_{t,j}$ is the probability of transmission per act modeled as a function of disease and care stage $\mathcal{s}_{t,j}$ and transmission risk group $\mathcal{r}_{j}$ of the infected node $j$; we will have a value of $\alpha_{j}=p_{t,j}$ if $j$ is a contact of $l$ (i.e., $V_{t}\left[ l,j \right]=1)$, is infected (i.e., $\mathcal{h}_{t,j}=1$), and is alive (i.e., $\mathcal{m}_{t,j}=1$), and $\alpha_{j}=0$ otherwise,

$\varepsilon$ = condom effectiveness,

$s_{t,j}$ = number of sex acts per month with node $j$, modeled as a function of age, transmission risk group, and number of partners of node $j$,

$c_{t,j}$ = proportion of acts with node $j$ that is condom protected, modeled as a function of age, transmission risk group, and number of partners of node $j$,

$F^{-1}\left( u \right)=$ an inverse Bernoulli distribution that takes a value of 1 with probability $u$ and value of 0 with probability $1-u$.

If node $l$ becomes infected, i.e., if the above equation yields $\mathcal{h}_{t,l}=1$, we set its HIV stage $\mathcal{s}_{t,l}=1$.

Every time-step $t$, this module also determines and updates any changes in sexual partnerships of HIV-infected nodes. Specifically, for every partnership $(k,j)$, it updates its active/inactive status as $V_{t}[k,j]=\left\{ \begin{aligned} 1\mathrm{if} \overline{t}\left( \left\{ k,j \right\} \right)\leq t\leq\underline{t}\left( \left\{ k,j \right\} \right), indicating it is active \\ 0 otherwise, indicating it is inactive \end{aligned} \right.$.

##### S1.3.3 ECNA network generation module for generating partnerships of new HIV-infected nodes

This module controls the overall network dynamics of partnerships between nodes. The main functionality is to generate the contact network for each new HIV-infected node $l$. The steps of the ECNA are as follows.

For every new HIV-infected node $l$,

1. Determine the number of new partnerships (edges) to generate as actual degree minus current degree. Note, these new partnerships would all be with HIV-susceptible persons as any partnerships with HIV-infected were already added when networks of those HIV-infected persons were created.
2. For each new HIV-susceptible partner node, determine node features: number of lifetime partners using *ECNA (discussed below)*, risk-group, current age-group, and pseudo-geographic jurisdiction using conditional probability distributions, and partnership distribution using a t*wo-step Markov process (discussed below)*.
3. For each partnership, determine age of both partners and simulation times at partnership initiation and termination, by formulating and solving as *assignment optimization model (discussed below).*
4. Determine who each new partner is by a uniform random draw from all who are eligible, i.e., all persons who are eligible have an equal chance of selection. All HIV-susceptible nodes in the network $G_{t}\mathcal{(N,E)}$ and HIV-susceptible non-agents in the compartmental model array $S_{t}$ with features matching that in steps 2 and 3 above, can be eligible.
5. For each new partner and partnership (determined in previous steps) update their corresponding features in the network $G_{t}\mathcal{(N,E)}$ and compartmental array $S_{t}$.

ECNA for determining degree of susceptible partner nodes

For a node $l$, the degree-bin of a partner $\bar{d}_{k}$ is not independent of its degree-bin $\bar{d}_{l}$ because of degree correlations between node neighbors, a key feature of scale-free networks [4], i.e., $\Pr\left( \bar{D}_{k}=\bar{d}_{k}|{\bar{D}_{l}=\bar{d}}_{l} \right)\neq\Pr\left( \bar{D}_{k}=\bar{d}_{k} \right); where \bar{D}_{k}\text{ is the random variable for degree-bin of node k}$, and thus, $\bar{d}_{k}$ cannot be directly drawn from its scale-free network probability mass function. While the literature presents an analytical method for estimation of $\Pr\left( \bar{D}_{k}=\bar{d}_{k}|{\bar{D}_{l}=\bar{d}}_{l} \right)$ for general static scale-free networks [4], this method is not suitable for simulating an epidemic in a dynamically evolving contagion network [1]. Specifically, in general static scale-free networks, the full network is available so the degree of all node neighbors is available, and thus $\Pr\left( \bar{D}_{k}=\bar{d}_{k}|\bar{D}_{l}=\bar{d}_{l} \right)$ is an expectation over all possible values of $\bar{d}_{l}$, i.e., an average over “all” node neighbors. However, as we only simulate HIV-infected nodes and their immediate contacts in the network, in our context, only values corresponding to HIV-infected node neighbors are used. As it is more likely that nodes with higher degree get infected first, the value of $\Pr\left( \bar{D}_{k}=\bar{d}_{k}|\bar{D}_{l}=\bar{d}_{l} \right)$ when $l$= ‘all node neighbors’ is different compared to when $l$= ‘HIV-infected node neighbors’. And further, $\Pr\left( \bar{D}_{k}=\bar{d}_{k}|\bar{D}_{l}=\bar{d}_{l} \right)$ is likely to change over time as the HIV epidemic spreads and the percent of the population that is HIV-infected changes. We developed a neural network model for the prediction of $\Pr\left( \bar{D}_{k}=\bar{d}_{k}|\bar{D}_{l}=\bar{d}_{l} \right)$ using as independent variables, $\bar{d}_{l}$, $\bar{d}_{k}$, the minimum degree of the network *m*, the percent of the population that is infected, and the scale-free network parameter $\lambda_{\mathcal{r}_{l}}$ corresponding to transmission risk group of node $l$ ($\mathcal{r}_{l})$. Details of this algorithm are presented in [1].

Two-step Markov process for determining the partnership distribution matrixfor each new partner

Suppose $d_{k}$ is the actual degree (number of lifetime partners) of a newly added susceptible node $k$. We need to determine at what age of $k$ will each partnership initiate. Specifically, suppose there is a matrix $\bar{L}$ of size $A\times D,$with $A$ the number of age-groups and $D$ the number of degree-bins, and element $\bar{L}[\bar{a},\bar{d}]$ representing the proportion of partnerships that initiate at age-group $\bar{a}$ for persons in degree-bin $\bar{d}$, with each column of $\bar{L}$ adding to 1, for all $\bar{d}\in\left\{ 1,2,\ldots D \right\}$. Then, for any node $k$ with actual degree $d_{k}\epsilon\bar{d}$, we can calculate the partnership distribution matrix, i.e., the number of partnerships that initiate at age $\bar{a}$, as $L_{k}\left[ \bar{a}, 1 \right]=\bar{L}\left[ \bar{a},\bar{d} \right].d_{k}$. Direct data for $\bar{L}$ would be a longitudinal survey over the duration of life of an individual, where the individual reports the number of partnerships they initiated at every age point of their life. Such surveys, however, are unavailable. Therefore, we estimated $\bar{L}$ using survey data on the reported number of partners up to that time by persons of different age-groups. Note that, these survey data only represent the number of partners up to the current age of the surveyed individual. Thus, the degree-bin $\bar{d}$ each person would belong to is unknown as $\bar{d}$ represents the number of partners the person would eventually have over their full lifetime. The age at which each partnership initiated is also unknown.

As data for $\bar{L}$ are not directly available, we developed a two-step estimation process. In step 1, we solved for the probabilities of initiating a new partnership when a person ages from age-group $\bar{a}-1$to $\bar{a}$, by formulating it as the transition probability matrix of a Markov chain and solving for it using an optimization model, calibrating to the survey data. In step 2, we used the transition probability matrix from step 1 to simulate partnership changes over the lifetime of a person, starting from age-group $\bar{a}=1 to age-group \bar{a}=A$, repeating for 10,000 people, grouping together all persons who end in the same degree-bin $\bar{d}$ at the last age-group $A$, and for each degree-bin $\bar{d}$, calculating the average proportion of partnerships that initiated in age-group $\bar{a}$, $\forall\bar{a}$. Details of this method are presented in [2].

Assignment model with heuristic solution for determining partnership initiation and termination age and time

We formulated the problem of assigning age at partnership initiation as a variant of an assignment optimization model. Suppose $n_{k}\left[ i \right]$ is the age of partnership initiation for $i^{th}$ partner of node $k$ (${|n}_{k}|=d_{k}$ the degree of node $k$). Then the objective is to assign each element (and all elements) of $n_{l}$ of a newly infected node $l$ to one (and-only-one) element of $n_{k}$ for each partner $k$, with constraints to maintain the probability distribution for age-mixing between partners, and partnership distribution matrices ($L_{t,l}$and $L_{t,k}, \forall k$) of the newly infected node $l$ and each partner $k$. However, solving it using standard solvers for each infected person is computationally expensive, and therefore, we developed a heuristic solution algorithm. Solving for partnership initiation age for each new partner sets the focal point for assigning the other three parameters partnership initiation time, partnership termination age, and partnership termination time. Details of this method are presented in [2].

##### S1.3.4 Disease progression module for simulating disease 1 progression for nodes in the network

The disease progression module updates the individual-level demographic and disease dynamics for every HIV-infected person in the network just like an agent-based model. This includes aging, Disease 1-related and natural mortality, progression through Disease 1 disease and care stages. For the analyses in the main paper, we adopted the disease progression module from PATH 2.0 [5].

### S2 HPV and cervical cancer (disease 2)

#### *S2.1 HPV natural history model*

The natural history of HPV and cervical cancer incidence and progression is presented in Figure S1 and was adapted from models in the literature [6–9]. For women, once a person is infected with HPV, the infection can persist, clear naturally by developing type-specific immunity or progress to cervical intraepithelial neoplasia grades I, II and III (CIN1, CIN2, CIN3); CIN 3 can then progress to invasive cervical cancer, progressing sequentially to localized, regional, and distant cancer stage. We only modeled cervical cancer that initiate as HPV infection. Persons diagnosed with cervical cancer, detected through either screening or natural symptoms, move to a diagnosed stage and face a certain chance of stage-specific mortality (the label in the figure corresponds to the stage at diagnoses). We stratified HPV types into three groups: HPV 16/18 pooled, HPV-31/33/45/52/58 pooled (high risk HPV types included in HPV 9 vaccine); other high-risk HPV pooled, thus we only modeled HPV-induced cervical carcinogenesis associated with all high-risk HPV types in this work. We did not model low-risk HPV types as their prevalence and risk of progression to cancer is low. We assume each HPV genotype group is independent of others, i.e., one cannot infect with multiple genotypes at the same time. We also assume infection-acquired immunity will wane over time while vaccine-acquired immunity stays life-long, and vaccine has 100% efficacy against corresponding HPV genotypes.

For MSM, we simulated HPV and anal cancer using the same epidemiology as HPV and cervical cancer [10]. For heterosexual males, we only modeled genotype-specific HPV infection and natural immunity (Figure S2). We first present the mathematical formulation of this natural history model, followed by data assumptions for incidence and progression.

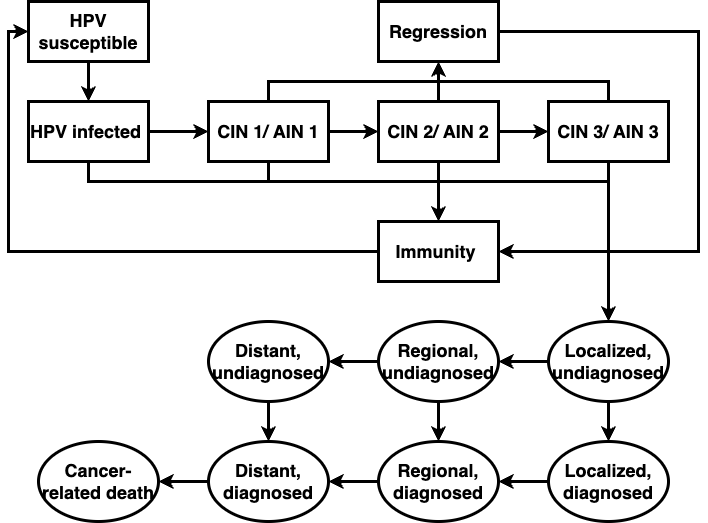

**Figure S1:** HPV and cancer disease diagram for HETF and MSM. Note: we assumed same epidemiology structure for HPV and anal cancer among MSM.

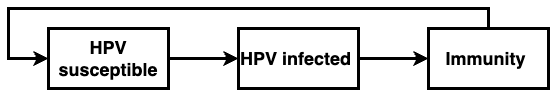

**Figure S2:** HPV natural history diagram for HETM.

#### *S2.2 Overview of compartmental model structure for hpv*

As noted in Section 1, the general equation for the compartmental model can be written as,

$S_{t}=S_{t-1}+\frac{{dS}_{t}}{dt}\Delta t$; $\frac{{dS}_{t}}{dt}=S_{t-1}G_{t}$

$\frac{{dS}_{t}}{dt}$ is the rate of change in $S_{t}$, modeled as Markov chain state transitions through non-stationary generator array $G_{t}$. The compartmental model array $S_{t}$ for HPV is of size $7\times3\times9\times1 \times41$, corresponding to size of age-group, risk-group, degree-bin, geographical jurisdiction, and HPV health state, and thus, $G_{t}$ is of size $7\times3\times9\times1 \times41\times41$. To simplify the notation, we define $v=[\bar{a}, \bar{r}, \bar{d},g]$, and rewrite the above equation as

$S_{t,v}=S_{t-1,v}+\frac{{dS}_{t,v}}{dt}\Delta t$; $\frac{{dS}_{t,v}}{dt}=S_{t-1,v}G_{t,v}$ such that $G_{t,v}$ is a $41\times41$ matrix, representing the Markov chain generator matrix for modeling transitions related to age-group $\bar{a}$, risk-group $\bar{r}$, degree-bin $\bar{d}$, and geographical jurisdiction $g$. Note, here we only have one jurisdiction, as it is a national model.

The 41 health states of the compartmental model include a disease-free state ($\bar{h}$ =0), and 14 disease states, $\bar{h}=s_{\tau}\in\{1,\ldots,14\}$ for each of the three HPV genotypes$\tau\in\{1,2,3\}$, such that ${\bar{h}=1}_{\tau}$ is the first stage of the disease for HPV genotype$\tau$. Thus, we can represent the force of infection as $G_{t,v}\left( 0, 1_{\tau} \right)=\lambda_{t,v}(0,1_{\tau})$, which we calculate using the method in Section 2.3. All other state transitions are as in a general Markov chain (see Section 1.3.1 for the simple two disease stage transition).

#### *S2.3 HPV transmission*

We simulated HPV transmission as a function of partnership acquisition and dissolution, by transmission risk group, age group, and degree bin. HPV infection can be transmitted, depending on the number of partner change per time unit, type-specific HPV prevalence among the group of partners, probabilities of HPV transmission per sex act, number of sex acts per time unit and number of partners per time unit as a proxy to duration of partnership. We present the mathematical formulation below.

##### S2.3.1 Force of infection

The overall force of infection $\lambda_{t,v}(0,1_{\tau})$ for persons in risk-group $\boldsymbol{v}$ at time $t$ for HPV type $\tau$, is calculated using

$$\lambda_{t,v}(0,1_{\tau})=\sum_{v} \bar{p}_{v^{'}v,\tau}\sum_{v'} [\bar{\Delta n_{vv^{'}}} \frac{I_{t,v^{'},\tau}}{I_{t,v^{'},\tau}+S_{t,v^{'}}}]$$

where,

$\bar{p}_{v^{'}v,\tau}$ is HPV transmission probability per sexual partnership from transmission risk group $v'$ to $v$ for HPV genotype $\tau$,

$\bar{\Delta n_{vv^{'}}}$ is the expected contact rate change for persons in transmission risk group v in initiating partnership with persons in transmission risk group v’ per unit of time,

$I_{t,v',\tau}$ is the number of people who are infected with HPV-type $\tau$ infection in transmission risk group $v'$at time step $t$,

$S_{t,v^{'}}$ is the number of people who are HPV negative in transmission risk group $v'$ at time step $t$.

##### S2.3.2 Transmission probability per partnership

As is common, the transmission probability per partnership is calculated by using a Bernoulli equation. We assume it is time-invariant

$$\bar{p}_{v^{'}v,\tau}=1-\left( 1-(1-\varepsilon)p_{v^{'}v,\tau} \right)^{ce_{v}}\left( 1-p_{v^{'}v,\tau} \right)^{\left( 1-c \right)e_{v}}$$

$$e_{v}=\frac{s_{v}}{n_{v}}$$

where,

$p_{v^{'},v,\tau}$ is the transmission probability per sex act from person in transmission risk group $v'$ to $v$ by HPV type $\tau$,

$e_{v}$ is the average number of exposure or average number of sexual acts per partnership for persons in transmission risk group $v$ per unit of time,

$s_{v}$is the number of sexual acts per time unit for persons in transmission risk group$v$,

$n_{v}$ is the number of sexual partners per time unit for persons in transmission risk group$v$,

$c$ is the proportion of condom protected acts per time unit for persons in transmission risk group$v$,

$\varepsilon$ is condom effectiveness, defined as the probability of reducing HPV transmission.

##### S2.3.3 Partnership mixing pattern

We calculate $\bar{\Delta n_{vv^{'}}}$, the number of partners per person for a person in risk-group$v$ with persons in risk-group $v'$ per time unit using time-invariant mixing matrices as follows

$$\bar{\Delta n_{vv^{'}}}=\Delta n_{v}\times\phi_{r}\left( a,a^{'} \right)\bigotimes\varphi_{r}\left( d,d^{'} \right)\bigotimes\psi_{r}\left( g,g^{'} \right)\bigotimes\chi\left( r,r^{'} \right)$$

where,

$\phi_{r}$ is the aging mixing matrix, element $\phi_{r}\left( a,a^{'} \right)$is the probability a person in age-group $a$ mixes with persons in age-group $a^{'}$ for transmission-group $r\in\{HETF, HETM, MSM\}$.

$\varphi_{r}$ is the degree bin mixing matrix, element $\varphi_{r}(d,d^{'})$ is the probability a person in degree bin $d$ form partnership with persons in degree bin $d'$ for transmission-group $r$.

$\psi_{r}$ is the jurisdiction mixing matrix, element $\psi_{r}(g,g^{'})$ is the probability a person in geographical location $g$ mixes with persons in geographical location $g^{'}$ for transmission-group $r$.

$\chi$is the transmission-group mixing matrix, element $\chi\left( r,r^{'} \right)$ is the probability a person in transmission-group $r$ forms partnership with a person in transmission-group $r'$.

#### *S2.4 Data input assumptions*

##### S2.4.1 Natural history parameters

The natural history model for stage transitions was adopted from [9]. Progression and regression rates for the stage transitions were adopted from [9,11,12]. Progression rates in cervical cancer stages, mortality rates by cancer stage, and probability of diagnoses by cancer stage can vary based on access to care and treatment (e.g., persons seeking care upon show of symptoms), so we took data specific to US from SEER database which is presented in [6]. Due to lack of data for anal cancer specific stage transition rates among general population, we assumed same rates as for cervical cancer.

##### S2.4.2 Sexual behavior parameters

To be consistent with the sexual behavior in the network model, we calculated or took the following parameters from the PATH 4.0 which were originally from [2,13–15]:

**Partnership change rate:** The partnership change rate was calculated using the following equation:

$$\frac{p_{d,r}\left( a \right)\times|d|}{\left| a \right|}$$

$p_{d,r}\left( a \right)$ represents the probability of partnership initiation at age group $a$ for people in degree bin $j$ and sexual transmission risk group $r$, which was calculated using the estimated proportion of partnerships initiated by age-group for persons with lifetime-partners in degree bin. $\left| d \right|$ is the number of lifetime partners in degree bin $d$ and we took an average of the degree bin range. For example, if $d$ represents degree bin $5 to 8$, then $\left| d \right|=\frac{5+8}{2}=6.5$. $\left| a \right|$ is the time interval representing the width of the age-group.

**Number of annual partners:** We used simulation techniques to estimate expected number of annual partners. We made following assumptions to ensure the output would be consistent with sexual behavior in the network model:

- An existing partnership without concurrency will end when the next partnership initiates or until one reaches the last age.

- An existing partnership with concurrency will end $\lambda$ unit of time after the next partnership initiates, where $\lambda$ follows an exponential distribution.

**Mixing matrix:** The age-group mixing matrix was calculated by observed data that was extracted from the report [16]. We assume sexual contact network follows scale-free network structures, thus given one’s degree, their neighbor’s degree can be calculated by using the degree correlation equation derived by [4]. The degree bin mixing matrix is then calculated by combining each individual degree together to logarithm-2 bin. The transmission risk group mixing matrix was obtained by assuming heterosexual males only have sex with heterosexual females and 20% of partners is heterosexual females among 29% of MSM who are bi-sexual [17–20].

**Other parameters:** All other parameters were either from the literature directly or from estimation and assumption presented in [2]. The data includes proportion of condom-protected acts for main and casual partnership, number of sex acts per year, condom effectiveness. We assume condom effectiveness for HPV is the same as that for HIV. We also assume in the compartment model, proportion of condom-protected acts for main partnership is applied to the group where annual partnership is less than or equal to one, otherwise proportion of condom-protected acts for casual partnership is applied.

##### S2.4.3 Demographic parameters

**Initialization:** We initialized the compartmental model to represent age-group distribution and risk-group distribution of people in the United States in 2006. We split the population by their HIV-state, age group, degree bin, sexual transmission risk group and geographical location. We initialized the number of people in each age and transmission risk group by distributing individuals to match with the US census data presented in [2]. The age group distribution does not sum to 1, since we only consider age from 13 to 100 in our model. We assume that 49.03% are heterosexual female, 48.70% as heterosexual male, and 2.27% of the US population are MSM.

**Births**: We modeled births in such way that individuals enter the compartment model by going into the youngest age group where people become sexually active. We assume people start sexually active as early as the age of 13 and hence there is no transmission for HIV and HPV prior to the youngest age in the model. Individuals are susceptible to both HPV and HIV upon their entering the population. We assumed a constant rate of entering the population α (overall birth rate in the US) and we used this rate multiplied by the number of individuals in the lowest age group (age 13 to17) of corresponding transmission risk group, degree bin and geographical location.

**Mortality:** Individuals leave the compartment model in two different ways: 1) Deaths and 2) aging out of the last age (age 100). Deaths in the compartmental model can result from: 1) cervical cancer 2) all-cause mortality (exclude HIV/AIDS and cervical cancer). We assume the oldest age for HIV and HPV transmission through sexual activity is 65 years, since the age of less than 2% of newly diagnosed HIV-infected individuals were 65 years and above. Mortality that contributes from cervical cancer varies by cancer stage, age and diagnosed status.

**Aging:** Individuals move on to the next age groups of corresponding transmission risk group, degree bin, geographical location and disease state from their current age group at a constant rate equal to inverse of the interval in age group a. Except for the youngest and oldest age group, all compartments experience inflows from the prior age group and outflows into the next age group. The youngest age group (age 13 to 17) receives inflow through births. The transitioning out of the last age group (age 65 to 100) results in their exiting the population.

#### *S2.5 Calibration and validation*

We calibrated the transmission probability per sex act to match epidemiological data targets used in the CISNET study [21], specifically, HPV prevalence and cervical cancer incidence and mortality from the era prior to implementation of HPV and cervical cancer screening. To do so we simulated the model to steady state by doing a dry-run for 70 years at 6-month time steps and repeating this with different probabilities of transmission until we found the set of values that gave a good fit to the following data metrics.

- Pre-screening genotype distribution among high-risk CIN 1, 2 and 3 from New Mexico HPV Pap Registry study [22].
- Pre-screening age- specific HPV prevalence among normal cytology from New Mexico HPV Pap Registry study [22].
- Pre-screening age-specific HPV genotype frequency among normal cytology obtained from New Mexico HPV Pap Registry study [22].

To validate the model, we compared our model outcome with cervical cancer incidence and mortality from the Connecticut Tumor Registry (CTR) for the period 1950-1969 [23] (See Figure 3a in the main manuscript). We also used the above calibrated model and screening rates from year 2006 in the U.S., to simulate HPV prevalence and cervical cancer incidence to steady state by doing a dry-run for 30 years at 6-month time steps. Specifically, we adopted age-specific cervical cancer screening utilization rate in 2006 [24], and screening sensitivity, proportions receiving follow-up colposcopy/biopsy and proportion receiving precancer treatment for those in pre-cancer stages, and additionally cancer-screening sensitivity for those in cancer stages as per [24,25]. Our model output is consistent with cervical cancer incidence and mortality reported more recently [26] (See Figure 3b in the main manuscript).

### S3 Model initialization and dry run

We can initialize the model to be representative of people in a population in a specific year. For the analyses in the main simulation, we initialized age, risk-group, degree distributions as per persons in the U.S in 2006 in both compartmental model and network. For distribution by Disease 2 health status in the compartmental model and network, we randomly selected 30% of persons in each category as infected with Disease 2, and dry running the simulations so that the state distributions reach a steady state. We initialized the network for Disease 1 to be representative of HIV in the US in the year 2006 through using two dry runs to ensure network dynamics are generated in addition to epidemic and demographic distributions.

Dry run is a technique used for initialization of the model. It involves running the simulation for a certain period of time, but the data generated over that period of run is not representative of an actual epidemic projection and thus referred to as a dry run. For a pure agent-based model (without networks), we could initialize the model with a few agents and assign parameters to match surveillance data for the demographic, disease stage, and care-continuum stage distributions. But the agents would be lacking the ‘history’, e.g., the age at infection, and the age and stage of ART initiation, relevant for modeling future events. Therefore, we can do a dry run, which means starting with some number of people, assigning them data according to a specific year, say 2006 surveillance distributions as done here for HIV, running the simulation for several years while maintaining the distributions to match that of 2006. As persons become newly infected, their history is being generated, and the initial persons from day 0 age-out of the model. The above method is also sufficient for diseases modeled in the compartmental model (Disease 2 here). However, for diseases modeled in ABENM (Disease 1 here), the network adds another layer of complexity as static data is infeasible (and moreover, rarely available) for determining the contact network structure, including the links of HIV-infected persons to each of their lifetime partners, the current age of partners, the initiation and termination times of the links, the initiation and termination age of nodes at both ends of a link, the risk-group of the partner, and the infection status of partner. Therefore, for Disease 1, we first do a dry run to allow for the network to dynamically grow over time (Dry run #1), and then apply the data from surveillance to set the demographic, and disease and care-continuum stages (Dry run #2). Dry runs are explained in more detail in [2] as applied to HIV and validated against NHSS, a similar method can be applied for other diseases modeled.

### S4 Overview of simulation modeling steps

We provide an overview of the full simulation model in this section. All notations used in the model are also summarized in Table S1.

Step 1: Initialization of the compartmental model array $S_{t=0}$ and graph $G_{t=0}\mathcal{(N,E)}$ at time-step $t=0$

Step 1a. We initialize the compartmental model array to be representative of the U.S. population by age, transmission risk group, and degree distribution in 2006. We distributed the total population in the U.S. into degree bins by using $Pr\left( 2^{\bar{d}-1}+1\leq k\leq2^{\bar{d}} \right)=\frac{\sum_{k=2^{i-1}+1:2^{i}} k^{-\lambda}}{\sum_{d=m:\bar{m}} d^{-\lambda}} \forall i=\{1,\ldots,7\}$, assuming minimum degree $m=2^{1}$ and maximum degree $\bar{m}=2^{7}$, and estimating the power-law parameter λ from life-time partner distribution data for MSM and heterosexuals [16]. Within each degree-bin, we distributed the population into seven age-groups, ranging from age 13 to 65, using US census data for distribution by age, and further by risk-group (heterosexual male, heterosexual female, or MSM) using population size estimates for MSM from [27,28].

Step 1b. We initiate a random graph of 1500 nodes and zero edges. For each node, we set their HIV status as newly infected, allocate a degree by randomly drawing from the overall degree distribution, and assign Disease 1-related care continuum and disease stages, age, and transmission risk group through random draws from the corresponding distributions of HIV in the US in 2006.

Step 2: Dry run to make the network and disease state-distribution representative of the population of interest

Dry run the model for a certain number of time-steps (here we chose 200 monthly time-steps) by running the following steps sequentially for every time-step.

Step 2a. Run the ECNA module (Section 1.3.3) to generate the network of contacts for every new HIV-infected node in the network.

Step 2b. Run the compartmental module (Section 1.3.1) to update Disease 2 parameters in the compartmental model and network.

Step 2c. Run the transmission module (Section 1.3.2) to generate new Disease 1 infections in the network.

Step 2d. Run the disease progression module (Section 1.3.4) to update demographics and Disease 1 progression and care parameters for nodes in the network.

Step 3: Main simulation run.

Repeat Steps 2a to 2d, for the required number of months. For the analyses in the paper, we simulated years 2006 to 2017 in monthly time-steps for Disease 1- and 6-month time-steps for Disease 2.

**Table S1: Table of Notations**

| **Notation** | **Description** |
| --- | --- |
| $t$ | Simulation time-step. |
| $A$ | The number of age-groups. |
| $R$ | The number of risk-groups. |
| $D$ | The number of degree bins. |
| $\mathcal{G}$ | The number of pseudo-geographic jurisdictions. |
| $H$ | The number of health states related to Disease 2. |
| $\bar{a}$; $\mathcal{r}$; $\bar{d}$; $g;$ $h$ | Used when referring to an age-group, risk-group, degree-bin, pseudo-geographic jurisdiction and health states, respectively. |
| $\mathcal{a}_{t,j}$;$\mathcal{r}_{j}$; $d_{j}$; $\mathcal{g}_{j};\mathcal{h}_{t,j}$ | Used when referring to the age, transmission-group, degree, geographic jurisdiction, and health status, respectively, of node $j$ in in the network at time $t$. Notations with no $t$ in subscript are variables whose values do no change over time. |
| $S_{t}[\bar{a}$, $\mathcal{r}$, $\bar{d}$, $g, h]$ | An array of size $A\times R\times D\times\mathcal{G\times}H$ representing the number of susceptible persons in the model, in age-group $\bar{a}$, transmission risk group $\mathcal{r}$, degree-bin $\bar{d}$, pseudo-geographic jurisdiction $g$, and health state $h$ at time t. |
| $\mathcal{N}$ | A set of nodes, each representing an infected person or a susceptible sexual partner. |
| $\mathcal{E}$ | A set of edges representing sexual partnerships between nodes. |
| $G_{t}\mathcal{(N,E)}$ | A dynamic graph with $\mathcal{N}$ a set of nodes and $\mathcal{E}$ a set of edges, at time t. |
| $Q_{t}$ | The number of nodes in graph *G*, at time *t*. |
| $C_{t}[i,j]$ | A static adjacency matrix of size $Q_{t}$ $\times Q_{t}$, with static element $C_{t}[i,j]=1$ if $i$ and $j$ are sexual partners anytime during their lifetime and $C_{t}[i,j]=$0 otherwise. |
| $V_{t}[i,j]$ | A dynamic adjacency matrix of size $Q_{t} \times Q_{t}$ , with element $V_{t}[i,j]=1$ if $i$ and $j$ are sexual partners during month $t$ and $V_{t}[i,j]=0$ otherwise. |
| $\mathcal{e=}\left\{ i,j \right\}$ | An edge in graph $G_{t}$ representing a sexual partnership between $nodes$ $i$ and $j$ |
| $\overline{t}(\left\{ i,j \right\})$ | The partnership initiation time; represents the simulation month for partnership initiation. |
| $\underline{t}(\left\{ i,j \right\})$ | The partnership termination time; represents the simulation month for partnership termination. |
| $\left\{ {\bar{\mathcal{a}}}_{i},{\bar{\mathcal{a}}}_{j} \right\}$ | The age of nodes $i$ and $j$ at the time of their partnership initiation. |
| $\left\{ {\underline{\mathcal{a}}}_{i},{\underline{\mathcal{a}}}_{j} \right\}$ | The age of nodes $i$ and $j$ at the time of their partnership termination. |
| $\bar{a}_{t,j}$ | Age-group of node $j$ at time $t$. |
| $\mathcal{a}_{t,j}$ | Age of node *j* at time *t.* |
| $\bar{d}_{j}$ | Degree-bin corresponding to the number of lifetime partners of node $j$. |
| $d_{j}$ | The actual number of lifetime sexual partners of node $j$. |
| $\hat{d}_{t,j}$ | The number of lifetime sexual partners of person $j$ who are already added as nodes in graph *G* at time *t*. For infected nodes $d_{j}=\hat{d}_{t,j}$ for susceptible nodes in *G,* $d_{j}\geq\hat{d}_{t,j}$ |
| $L_{t,j}$ | A partnership distribution matrix of size $A\times2$, where $L_{t,j}[\bar{a},1]$ is the number of partnerships that initiate at age-group $\bar{a}$, and $L_{t,j}[\bar{a},2]$ is the number of partnerships that are yet to be assigned. For infected nodes,$L_{t,j}\left[ \bar{a},2 \right]=0,\forall\bar{a}$ |
| $\mathcal{h}_{t,j}$ | Infection status of node *j* at time *t*. |
| $\mathcal{m}_{t,j}$ | Deceased status of node *j* at time *t*. |
| $\mathcal{r}_{j}$ | Risk-group of person *j*. |
| $\mathcal{s}_{t,j}$ | Care continuum or disease stage of person *j* at time *t*. |
| $p_{t,j}$ | Infectiousness or risk of transmission per act for person *j* at time *t*. |
| $\varepsilon$ | Condom effectiveness. |
| $s_{t,j}$ | The number of sex acts per month for person *j* at time *t*. |
| $c_{t,j}$ | The proportion of acts condom protected of person *j* at time *t*. |
| $F^{-1}\left( u \right)$ | The inverse Bernoulli distribution that takes values 1 with probability $u$ and 0 with probability $1-u$. |
| $D_{k}$ | Random variable for degree of node $k$. |
| $Pr(D_{k}=d_{k}\vert{D_{l}=d}_{l})$ | Conditional probability distribution for $D_{k}$. |
| $Pr({D_{k}=d}_{k})$ | Marginal probability distribution for $D_{k}$. |
| *m* | Minimum degree of the network. |
| $\lambda_{\mathcal{r}_{l}}$ | Scale-free network parameter corresponding to the risk-group of node *l*. |
| $\bar{L}[\bar{a},\bar{d}]$ | A matrix of size $A\times D$, representing the proportion of partnerships that initiate at age-group $\bar{a}$ for persons in degree-bin $\bar{d}$. |
| $X_{\bar{a}}$ | Random variable representing the number of lifetime partners at age-group$\bar{a}$. |
| $P_{\bar{a}}$ | The transition probability matrix of size $D\times D$ for age-group $\bar{a}$. |
| $\pi_{\bar{a}}$ | The steady state distribution for lifetime-partners in age-group$\bar{a}$. |
| $z$ | A vector of size $D^{2}\times1$, converted from matrix $P_{\bar{a}}$. |
| $C$ | A matrix of size $D\times D^{2}$, recreated with values of $\pi_{\bar{a}}$. |
| $B$ | A binary matrix of size $D\times D^{2}$. |
| $b$ | A vector of ones of size $D\times1$. |
| $M[i,j]$ | An age-mixing matrix of size $A\times A$, which represents the probability that, given a person is in age-group$i$, his or her partner is in age-group $j$. Varies by risk-group. |
| $P\left[ i,j \right]$ | A matrix of size $A\times A$, representing the expected number of contacts the newly infected node $l$ should have with person in age-group $i$, when node $l$ is in age-group $j$. |
| $N\left[ \bar{a},k \right]$ | A binary matrix of size $A\times(d_{l}-\hat{d}_{t,l})$, which represents whether partner $k$ would have a newly initiating partnership at age-group $\bar{a}$. |
| $R\left[ \bar{a},k \right]$ | A binary matrix of size $A\times(d_{l}-\hat{d}_{t,l})$, which represents whether the partnership with partner $k$ would occur when partner $k$ is in age-group $\bar{a}$. |
| $n\left[ i \right]$ | A vector of size *A*, denoting the number of partnerships that should initiate when the partner is in age-group *i*. |
| $e_{k}[i]$ | A vector of size *A*, representing node *k* to be eligible to initiate a partnership at age-group *i*. |

**Table S2: Increased risk in HPV progression among HIV positive versus HIV negative persons (noted were applied to women and MSM, and heterosexual men where applies)**

| **Parameter** | **Stratification** | **Value** | **Range (LB)** | **Range (UB)** | **Reference** |
| --- | --- | --- | --- | --- | --- |
| HPV acquisition multiplier | By CD4 count |  |  |  | [12] |
|  | >500 |  | 1.7 | 2 |  |
|  | 350 - 500 |  | 1.9 | 2.1 |  |
|  | 200 - 350 |  | 2.1 | 2.3 |  |
|  | <200 |  | 2.2 | 2.5 |  |
| HPV to CIN 1 multiplier |  | 3.73 | 2.62 | 5.32 | [29] |
| CIN1 to CIN2 multiplier | By CD4 count |  |  |  | [29,30] |
|  | >500 | 1 |  |  |  |
|  | 350 - 500 | 1.75 |  |  |  |
|  | 200 - 350 | 2.3 |  |  |  |
|  | <200 | 2.66 |  |  |  |
| CIN2 to CIN3 multiplier | By CD4 count |  |  |  | [29,30] |
|  | >500 | 1 |  |  |  |
|  | 350 - 500 | 1.75 |  |  |  |
|  | 200 - 350 | 2.3 |  |  |  |
|  | <200 | 2.66 |  |  |  |
| CIN3 to cancer multiplier |  | 2.5 | 2.3 | 2.5 | [29,30] |
| Natural immunity clearance multiplier | By CD4 count |  |  |  | [12] |
|  | >500 | 6.8333 |  |  |  |
|  | 350 - 500 | 7.8664 |  |  |  |
|  | 200 - 350 | 9.4444 |  |  |  |
|  | <200 | 14.167 |  |  |  |
